# Does symptom perception after negative affect induction differ between physically ill and healthy individuals? An experimental study within SOMA.CK

**DOI:** 10.64898/2025.11.27.25341154

**Authors:** Birte Jessen, Christian Schmidt-Lauber, Tobias B. Huber, Bernd Löwe, Omer Van den Bergh, Michael Witthöft, Meike Shedden-Mora

**Author notes:** Correspondence to: Birte Jessen, Department of Psychology, Medical School Hamburg, Hamburg, Germany, Am Kaiserkai 1, 20457 Hamburg, Germany.

## Abstract

**Introduction:** Symptom perception is highly subjective and influenced by the complex interplay of biopsychosocial factors. This study aimed to explore whether negative affect induction using the Affect and Symptoms Paradigm (ASP) influences symptom perception in patients with non-dialysis chronic kidney disease (ND-CKD) and healthy controls.

**Methods:** Participants watched three picture series with varying affective content (positive, negative, neutral) from the International Affective Picture System (IAPS). After each picture set, participants rated symptom levels (10-item symptom checklist), affective state (Positive and Negative Affect Schedule) and arousal levels (Self-Assessment-Manikin-System).

**Results:** In N = 115 individuals with ND-CKD from the SOMA.CK study (mean age = 62.95, SD = 12.60) and 100 age- and gender-matched healthy controls (mean age = 60.00, SD = 12.80) negative pictures significantly increased negative affectivity and arousal. All participants reported significantly higher symptom levels after negative versus positive and neutral pictures. No significant group differences in overall symptom levels emerged. Habitual symptoms did not moderate symptom levels after negative affect induction, although high habitual symptom reporters showed higher symptom levels across all picture categories. In the CKD group symptom levels were moderated by difficulties in identifying feelings and in controls by suppression as emotion regulation strategy.

**Conclusion:** Negative affect induction increases symptom levels, even in a chronic illness such as CKD. These results are in line with the predictive processing model which suggests that symptom perception develops from a complex inferential process of somatosensory input in light of pre-existing symptom representations in memory (priors).

**Highlights:**

- Patients with ND-CKD and healthy controls reported higher symptom levels after negative pictures
- Habitual symptoms did not moderate symptom levels after negative pictures
- Difficulties in identifying feelings moderated symptom levels in the CKD group
- Suppression moderated symptom levels in the HC group
- Symptoms after negative pictures predicted CKD-specific symptom burden at 6 months

## Introduction

How do individuals perceive and interpret own bodily sensations, and what factors shape their symptom perception? Symptom perception is highly subjective and influenced by many factors. Biological processes alone do not sufficiently explain symptom development [1]. Complex biopsychosocial interactions between psychological factors such as negative affectivity (NA), dysfunctional cognitions or biological factors such as inflammation contribute to symptom development and perception [2, 3]. *Persistent somatic* or *physical symptoms* (PSS/PPS) are defined as somatic symptoms with or without medical explanation being present on most days for at least several months [2, 4]. PSS are not only common in the general population but also in chronic conditions such as chronic kidney disease (CKD) [2].

According to the predictive processing model (PPM), the brain interprets somatosensory input based on pre-existing information that act as implicit predictions (*priors)* [5]. Depending on the balance between priors and somatic input, symptom perception can vary substantially between and within individuals [6, 7]. If the prior is very precise while the somatic input is less, the actual symptom perception will be more influenced by priors than by somatosensory input [6]. Conversely, highly precise somatic input will typically outweigh the impact of priors. High habitual symptoms, a negative affective state and maladaptive emotion regulation may impact symptom perception through their effect on priors [1, 3, 8–10].

Evidence shows that physical symptoms can reliably be induced by negative affect using affective picture viewing (Affect and Symptoms Paradigm, ASP) [11]. The ASP has primarily been used in individuals with functional somatic syndromes (FSS) or in healthy individuals, where watching negative pictures increased symptom reporting, especially in those with high habitual symptoms [9, 10, 12–14]. High habitual symptoms, emotion regulation deficits, trait NA, alexithymia and somatosensory amplification have been investigated as moderating factors of symptom perception [12]. Habitual symptoms moderated symptom levels after negative affect induction in former studies [13, 14]. Deficits in emotion regulation are known as potential moderating factors of symptom perception, especially in individuals with PSS [15]. High trait NA was associated with habitual symptoms [9, 16]. Alexithymia, in particular difficulty identifying feelings, moderated symptom levels after negative pictures [12]. Somatosensory amplification as process of increasingly perceiving bodily sensations and falsely interpreting those as danger due to hyper focusing was further identified as moderator [17]. A fMRI study in patients with FSS showed that elevated symptoms in the ASP were partly mediated by increased activation in somatosensory and pain processing brain patterns, indicating that affect-driven nociceptive priors are a key mechanism in FSS [18]. What remains unclear is if the existence of a chronic medical condition plays a specific role in symptom perception after negative affect induction and leads to more or less symptoms in the ASP. CKD is employed as model condition exploring this question, affecting 9.1% globally and often accompanied with cardiovascular disease or diabetes [19–21]. PSS in CKD, present in all disease stages, [19] affect quality of life, morbidity and mortality [21, 22]. Because high somatic symptom burden cannot fully be explained by CKD stage alone [19, 21, 23], the question is whether activation of priors in the brain impact symptom perception in CKD. Because the ASP assesses the impact of symptom priors relative to the impact of somatic input on the experience of symptoms, it is well suited to investigate this issue. Fatigue, pruritus or pain and additional symptoms in more advanced disease stages [23–25] may rely on high somatosensory input, and therefore lead to less symptoms after affect induction in the ASP compared to healthy controls. Conversely, persistent symptom experience could contribute to the development of strong, precise symptom priors and increase individuals’ vulnerability to report elevated symptoms in the ASP [12]. Very few studies have experimentally examined mechanisms of symptom perception in chronic symptomatic illnesses like CKD. Research on the ASP mostly investigated high habitual symptom reporters among healthy participants or individuals with FSS [10, 12, 14]. Therefore, the present study provides valuable insights to better understand mechanisms and moderating factors influencing symptom perception in chronic symptomatic illness (CKD) and healthy individuals.

We hypothesized that inducing negative affect increases symptom perception in patients with CKD as well as in healthy controls. We further explored whether patients with CKD and HCs experience comparable symptoms after watching negative pictures. In both groups, we expected this effect to be more pronounced in individuals with high habitual symptoms, deficits in emotion regulation, higher trait NA, alexithymia, and high levels of somatosensory amplification. In patients with CKD, we further explored the impact of disease severity (CKD stage) on symptom perception. Finally, we examined whether higher symptom levels after negative affect induction predicts greater general and CKD-specific symptom burden at 6 and 12 months.

## Methods and analyses

### Study design

This study is embedded in the longitudinal mixed-methods SOMA.CK study [26], a subproject of the SOMACROSS Research Unit (RU 5211), which investigates factors and mechanisms of somatic symptom persistence across diseases [2]. SOMA.CK investigates predictors of somatic symptom persistence in patients with CKD over 12 months [26]. The study protocol was approved by the ethics committee of the State of Hamburg Chamber of Medical Practitioners (reference number 2020-10195-BO-ff) and was preregistered on ISRCTN (ISRTCN16137374).

### Participants

An a-priori power analysis for a repeated-measures ANOVA, within-between interaction with an expected effect size *f* of 0.11, an α error of 0.05 and a power of 0.95 revealed 108 participants per group would be sufficient [12].

This cross-sectional study included 115 participants with CKD from SOMA.CK collaborating clinics and 100 healthy controls, matched by age and sex. Controls were recruited via posters in pharmacies, supermarkets, bus stops and one assisted living facility. Inclusion criteria for both groups were legal age and sufficient knowledge of German. Exclusion criteria included having cognitive impairment, schizophrenia, substance dependency or acute suicidality. Additional CKD-specific exclusion criteria are documented in the SOMA.CK study protocol [26]. Longitudinal data was used addressing the third hypothesis in the CKD group.

### Procedure

Individuals with CKD participated during their SOMA.CK study appointment, controls were assisted by study staff online or on-site. After informed consent, participants completed questionnaires. As recommended by the Center for the Study of Emotion and Attention at the University of Florida [11], participants were informed that negative pictures could temporarily evoke unpleasant emotions. Three sets of 20 pictures (8 seconds each) in permuted randomized order (figure 1) were presented. During debriefing, participants were given the opportunity talking to a psychologist.

**Figure 1:**
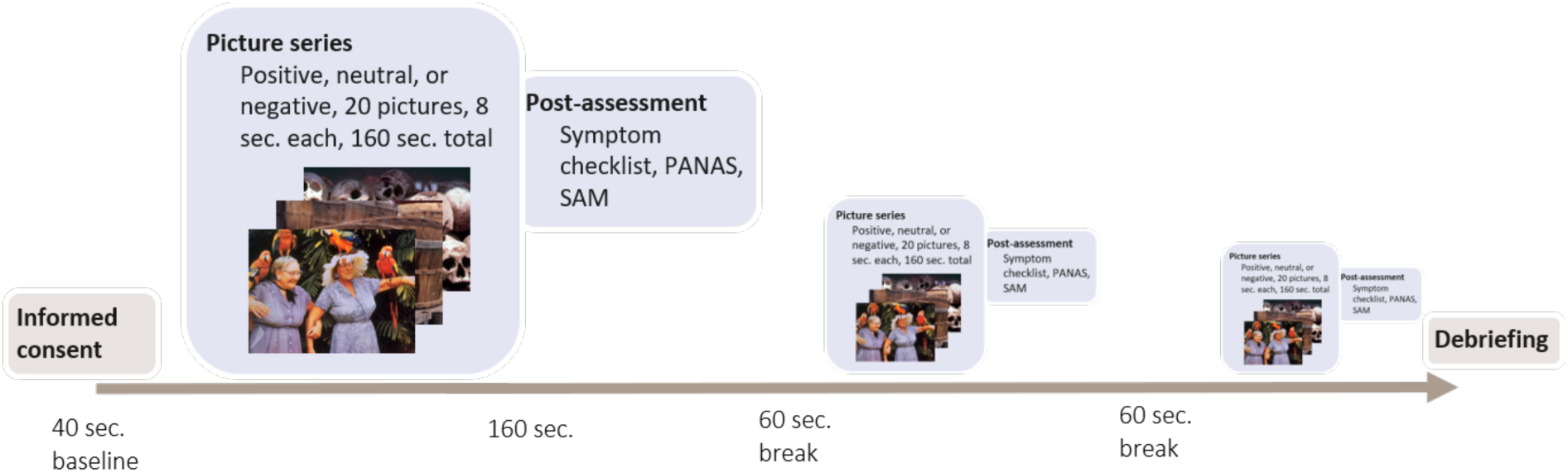
Experimental design of the Affective Picture Paradigm. Note. PANAS = Positive and Negative Affect Schedule; SAM = Self-Assessment Manikin assessing affective valence, arousal and control.

### Assessments and study outcomes

#### Instruments

The *Affect and Symptoms Paradigm (ASP)* assessed symptom perception after negative affect induction using 60 pictures from the *International Affective Picture System* (IAPS), a normative set of coloured pictures with positive, negative and neutral affect [11]. Based on [12], low disgust pictures were chosen, balanced by content and perspective (S1). The experiment was programmed in Tivian [27].

##### Primary Outcome

*Symptom levels* were measured with a 10-item-symptom-checklist [9, 12, 14] assessing 10 symptoms (tightness in the chest, heart pounding, abdominal or stomach pain or cramps, headache, tiredness or exhaustion, shortness of breath, heart racing, nausea, dizziness, muscle pain) on a five-point scale (*(1) not at all* to *(5) very strong)*. Sum scores ranged from 10-50, with higher scores indicating greater symptom levels. Additionally, the *ASP* effect (difference in symptom levels after negative versus neutral pictures) was calculated [10].

#### Manipulation checks

##### Valence, arousal and control

The Self-Assessment-Manikin (SAM) as language-free pictogram-based instrument assessed *valence* (pleasantness), *arousal* and perceived *control* after each picture set. Based on [14, 28], the nine-level scale was chosen.

##### Affectivity state

The Positive and Negative Affect Schedule state (PANAS state) measured current affectivity with 20 items (10 positive, 10 negative) on a five-point scale (*(1) not at all* to *(5) very much*) and mean values of positive (PA) and negative affectivity (NA) [29].

#### Moderators

##### Disease severity

CKD stages based on the glomerular filtration rate (GFR) according to guidelines [19] were calculated in the CKD group.

##### Habitual symptom burden

The Patient Health Questionnaire-15 (PHQ-15) assessed the presence and burden of 15 somatic symptoms during the past four weeks on a three-point scale (0 *not bothered at all*, 1 *lightly bothered*, 2 *bothered a lot)* with sum scores (0-30) and categories of *low* (0-4), *medium* (5-9) and *high* (≥10) [30].

##### Trait affectivity

The PANAS trait measured general affectivity over the past 12 months (see PANAS state) [29].

##### Alexithymia

The Toronto Alexithymia Scale 20 (TAS-20) assessed the *difficulty identifying feelings* (DIF) in one subscale, rated on a five-point scale (*(1) strongly disagree* to *(5) strongly agree*). A total score was calculated [31, 32].

##### Emotion regulation

The Emotion Regulation Questionnaire (ERQ) assessed two emotion regulation strategies [33]: *cognitive reappraisal* (6 items) and *expressive suppression* (4 items) rated on a seven-point scale (*(1) strongly disagree* to *(7) strongly agree*) and higher scores representing greater strategy use.

##### Somatosensory amplification

The Somatosensory Amplification Scale (SSAS) measured the tendency perceiving normal bodily sensations as intense or disturbing. Consisting of 10 items on a five-point scale (*(1) not at all* to *(5) extremely),* a total score was calculated [34].

#### Outcomes at 6 and 12 months

*Habitual symptom burden* at 6 and 12 months was measured with the PHQ-15 (see above) [30, 35].

*CKD-specific symptom burden* was measured using the Chronic Kidney Disease Symptom Burden Index (CKD-SBI) assessing *prevalence, distress, severity* and *frequency* of 32 symptoms on an eleven-point scale (*(0) none/never* to *(10) very much/very severe/constant*). A total score (0-100) was calculated by summing subscales and multiplying by 0.1008 [36, 37].

#### Data analysis

Missing analyses revealed 27% missing GFR values due to missing blood samples. CKD stages were added via medical records, enabling 110/115 participants with CKD in moderation analyses. One participant per group missed single questionnaires (PHQ-15, TAS-20, PANAS trait) and was excluded from moderation analyses. T-tests, chi-square tests, and bivariate correlations compared descriptive statistics. Repeated-measures ANOVAs (Greenhouse-Geisser corrected) examined manipulation checks and main analyses of affect induction on symptom levels within each group (CKD and HC), followed by planned contrasts between negative versus positive/neutral pictures. Further two-factorial ANOVA tested if group affiliation (CKD vs. HC) moderated symptom levels after affect induction using *affect induction* as within-subject and *group* as between-subject factor. Repeated measures ANOVAs with affect induction as within-subject and moderators as between-subject factor tested effects of moderators on symptom levels within each group. Median splits per group and (extreme) group comparisons tested moderation effects of disease severity (CKD stage), habitual symptom burden (PHQ-15: low, medium, high), trait NA (PANAS: median split), DIF (TAS-20: average ± 1SD), emotion regulation strategies (ERQ: median split) and somatosensory amplification (SSAS: median split). Correlations between the ASP effect and moderators were additionally calculated. Differences in moderators across groups (CKD vs. HC) were compared using two-factorial repeated-measures ANOVAs with affect induction as within-subject factor and group and moderators as between-subject factors. Predictive analyses of general and CKD-specific symptom burden at 6 and 12 months used bivariate correlations and multiple linear regression with symptom levels after negative affect induction and the ASP effect as predictor. Statistical analyses were conducted with SPSS 27.

## Results

### Sample characteristics

Table 1 shows demographic and clinical characteristics of the overall sample. Matching was successful with no group differences regarding age and sex. The CKD group had a significantly lower education, higher BMI, more medication use, higher habitual symptom burden, and lower trait PA.

**Table 1:**
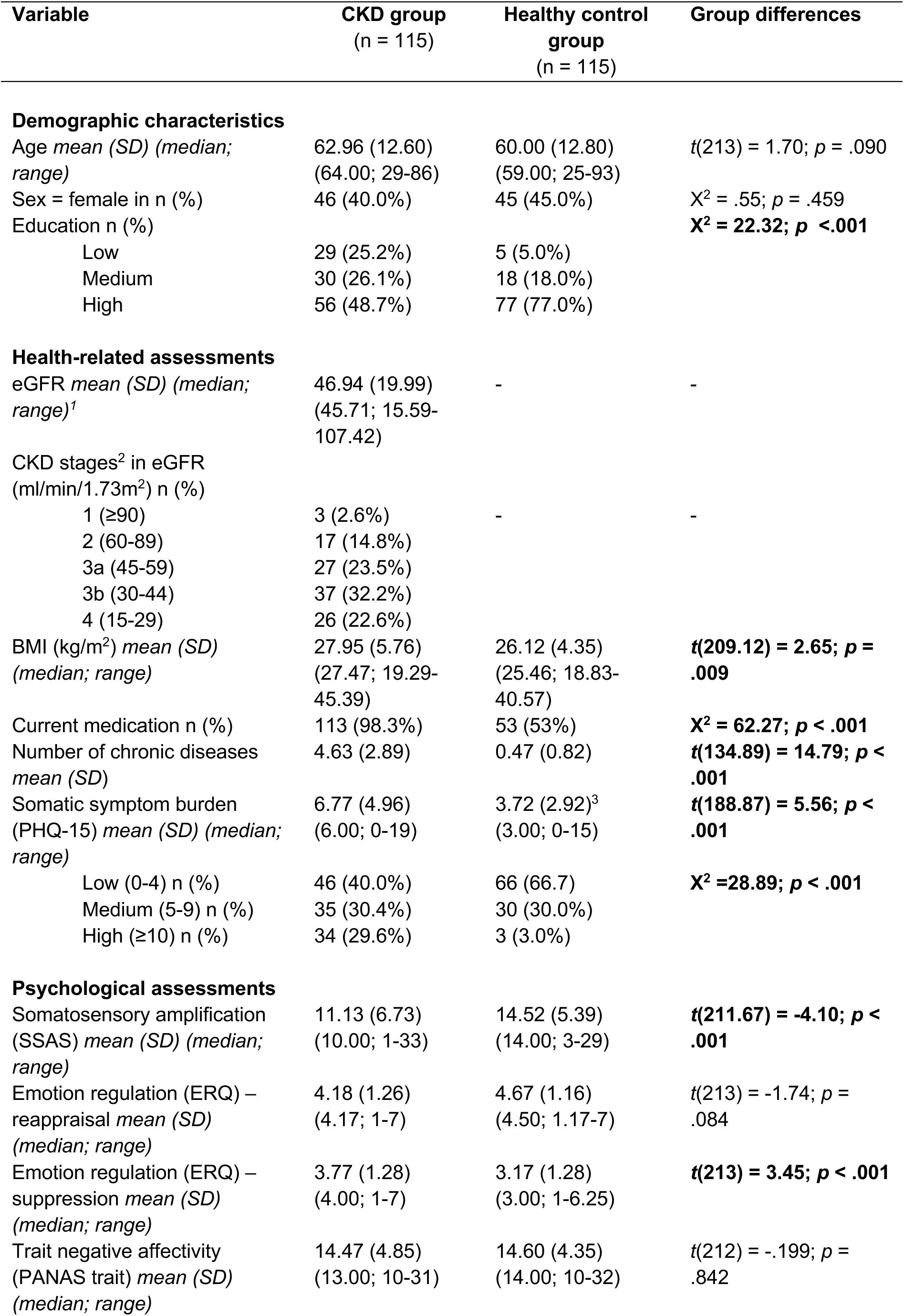
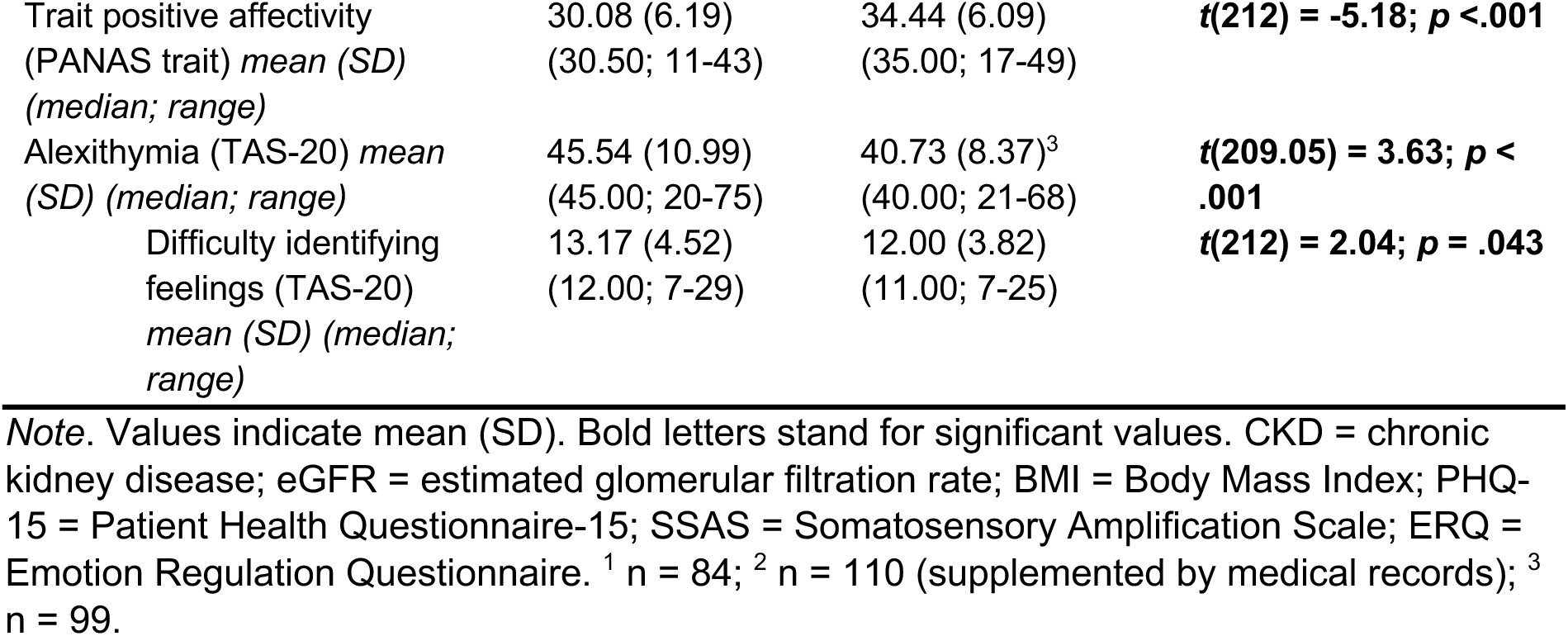
Demographic and clinical characteristics of participants.

| Variable | CKD group<br>(n = 115) | Healthy control<br>group<br>(n = 115) | Group differences |
| --- | --- | --- | --- |
| **Demographic characteristics** |  |  |  |
| Age *mean (SD) (median; range)* | 62.96 (12.60)<br>(64.00; 29-86) | 60.00 (12.80)<br>(59.00; 25-93) | $t(213) = 1.70; p = .090$ |
| Sex = female in n (%) | 46 (40.0%) | 45 (45.0%) | $X^2 = .55; p = .459$ |
| Education n (%) | | | $X^2 = 22.32; p < .001$ |
| Low | 29 (25.2%) | 5 (5.0%) |  |
| Medium | 30 (26.1%) | 18 (18.0%) |  |
| High | 56 (48.7%) | 77 (77.0%) |  |
| **Health-related assessments** |  |  |  |
| eGFR *mean (SD) (median; range)*<sup>1</sup> | 46.94 (19.99)<br>(45.71; 15.59-107.42) | - | - |
| CKD stages<sup>2</sup> in eGFR<br>(ml/min/1.73m<sup>2</sup>) n (%) |  |  |  |
| 1 ($\geq 90$) | 3 (2.6%) | - | - |
| 2 (60-89) | 17 (14.8%) |  |  |
| 3a (45-59) | 27 (23.5%) |  |  |
| 3b (30-44) | 37 (32.2%) |  |  |
| 4 (15-29) | 26 (22.6%) |  |  |
| BMI (kg/m<sup>2</sup>) *mean (SD) (median; range)* | 27.95 (5.76)<br>(27.47; 19.29-45.39) | 26.12 (4.35)<br>(25.46; 18.83-40.57) | $t(209.12) = 2.65; p = .009$ |
| Current medication n (%) | 113 (98.3%) | 53 (53%) | $X^2 = 62.27; p < .001$ |
| Number of chronic diseases<br>*mean (SD)* | 4.63 (2.89) | 0.47 (0.82) | $t(134.89) = 14.79; p < .001$ |
| Somatic symptom burden<br>(PHQ-15) *mean (SD) (median; range)* | 6.77 (4.96)<br>(6.00; 0-19) | 3.72 (2.92)<sup>3</sup><br>(3.00; 0-15) | $t(188.87) = 5.56; p < .001$ |
| Low (0-4) n (%) | 46 (40.0%) | 66 (66.7) | $X^2 = 28.89; p < .001$ |
| Medium (5-9) n (%) | 35 (30.4%) | 30 (30.0%) |  |
| High ($\geq 10$) n (%) | 34 (29.6%) | 3 (3.0%) | |
| **Psychological assessments** |  |  |  |
| Somatosensory amplification<br>(SSAS) *mean (SD) (median; range)* | 11.13 (6.73)<br>(10.00; 1-33) | 14.52 (5.39)<br>(14.00; 3-29) | $t(211.67) = -4.10; p < .001$ |
| Emotion regulation (ERQ) –<br>reappraisal *mean (SD) (median; range)* | 4.18 (1.26)<br>(4.17; 1-7) | 4.67 (1.16)<br>(4.50; 1.17-7) | $t(213) = -1.74; p = .084$ |
| Emotion regulation (ERQ) –<br>suppression *mean (SD) (median; range)* | 3.77 (1.28)<br>(4.00; 1-7) | 3.17 (1.28)<br>(3.00; 1-6.25) | $t(213) = 3.45; p < .001$ |
| Trait negative affectivity<br>(PANAS trait) *mean (SD) (median; range)* | 14.47 (4.85)<br>(13.00; 10-31) | 14.60 (4.35)<br>(14.00; 10-32) | $t(212) = -.199; p = .842$ |
| Trait positive affectivity<br>(PANAS trait) *mean (SD)*<br>(*median; range*) | 30.08 (6.19)<br>(30.50; 11-43) | 34.44 (6.09)<br>(35.00; 17-49) | ***t*(212) = -5.18; *p* < .001** |
| Alexithymia (TAS-20) *mean*<br>(*SD*) (*median; range*) | 45.54 (10.99)<br>(45.00; 20-75) | 40.73 (8.37)<sup>3</sup><br>(40.00; 21-68) | ***t*(209.05) = 3.63; *p* < .001** |
| Difficulty identifying<br>feelings (TAS-20)<br>*mean (SD) (median; range)* | 13.17 (4.52)<br>(12.00; 7-29) | 12.00 (3.82)<br>(11.00; 7-25) | ***t*(212) = 2.04; *p* = .043** |

*Note.* Values indicate mean (SD). Bold letters stand for significant values. CKD = chronic kidney disease; eGFR = estimated glomerular filtration rate; BMI = Body Mass Index; PHQ-15 = Patient Health Questionnaire-15; SSAS = Somatosensory Amplification Scale; ERQ = Emotion Regulation Questionnaire. <sup>1</sup> *n* = 84; <sup>2</sup> *n* = 110 (supplemented by medical records); <sup>3</sup> *n* = 99.

### Affect induction (manipulation check)

To determine whether affect induction was effective, we compared valence, arousal and control (SAM), and positive and negative affect (PANAS) after the three-picture series within each group. Then, we analysed if the groups differed in their response to the pictures.

Significant main effects for picture category of SAM and PANAS ratings were found within groups. Valence was significantly higher after positive compared to negative and neutral pictures. Arousal was significantly higher after negative than positive and neutral pictures. Perceived control was significantly lower after negative than positive and neutral pictures.

Positive affectivity significantly differed across all picture series, peaking after positive pictures. Negative affectivity differed across all picture series in the CKD group and was higher after negative than positive and neutral pictures in the HC group.

Comparisons between the CKD and the HC group showed only few main effects in manipulation checks. Significant main and interaction effects for positive affectivity indicated higher ratings in controls after positive pictures. Further interactions showed controls rated positive pictures as more pleasant, negative as more unpleasant, felt more aroused, and perceived less control after negative pictures than the CKD group (Table 2).

**Table 2:**
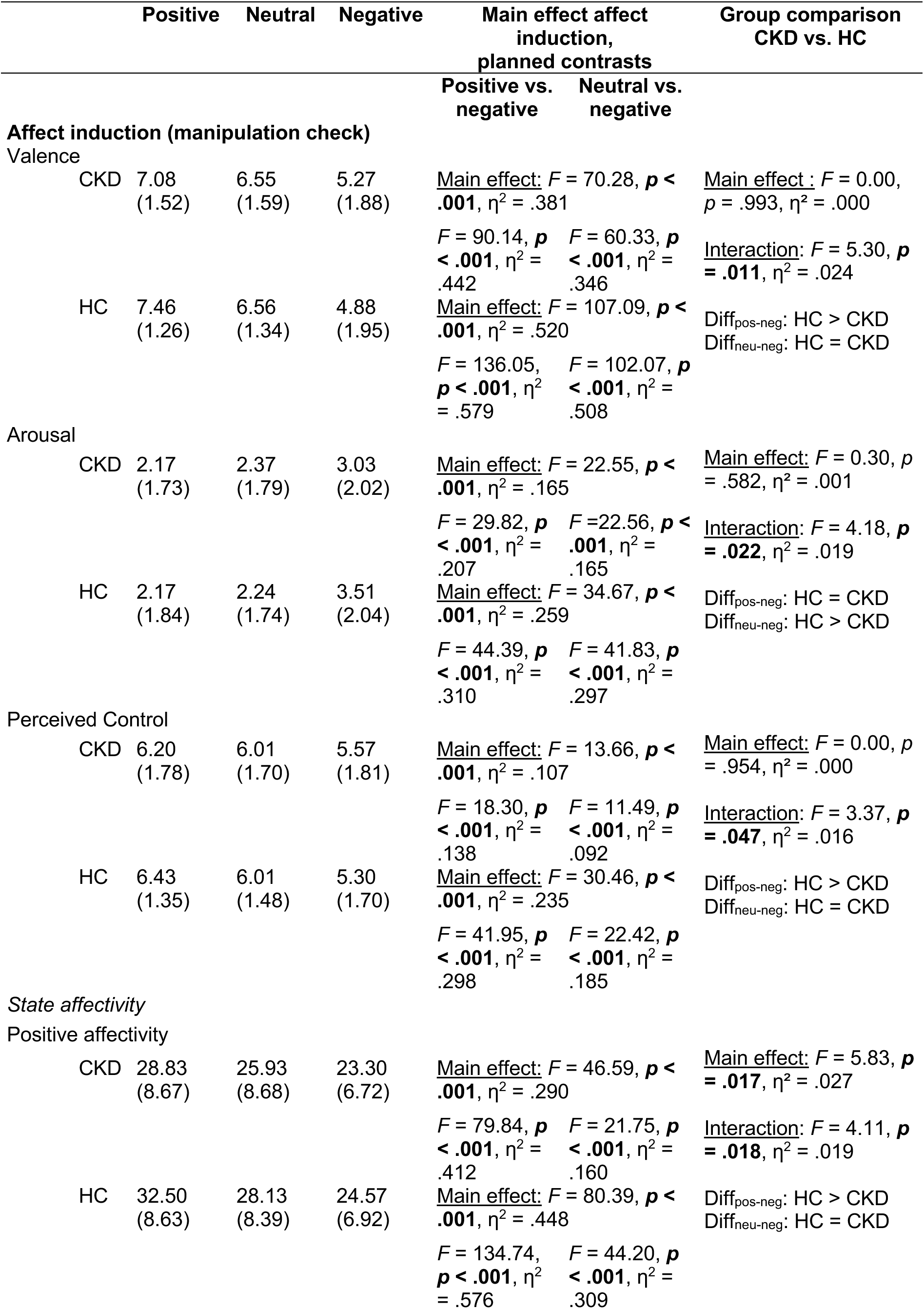
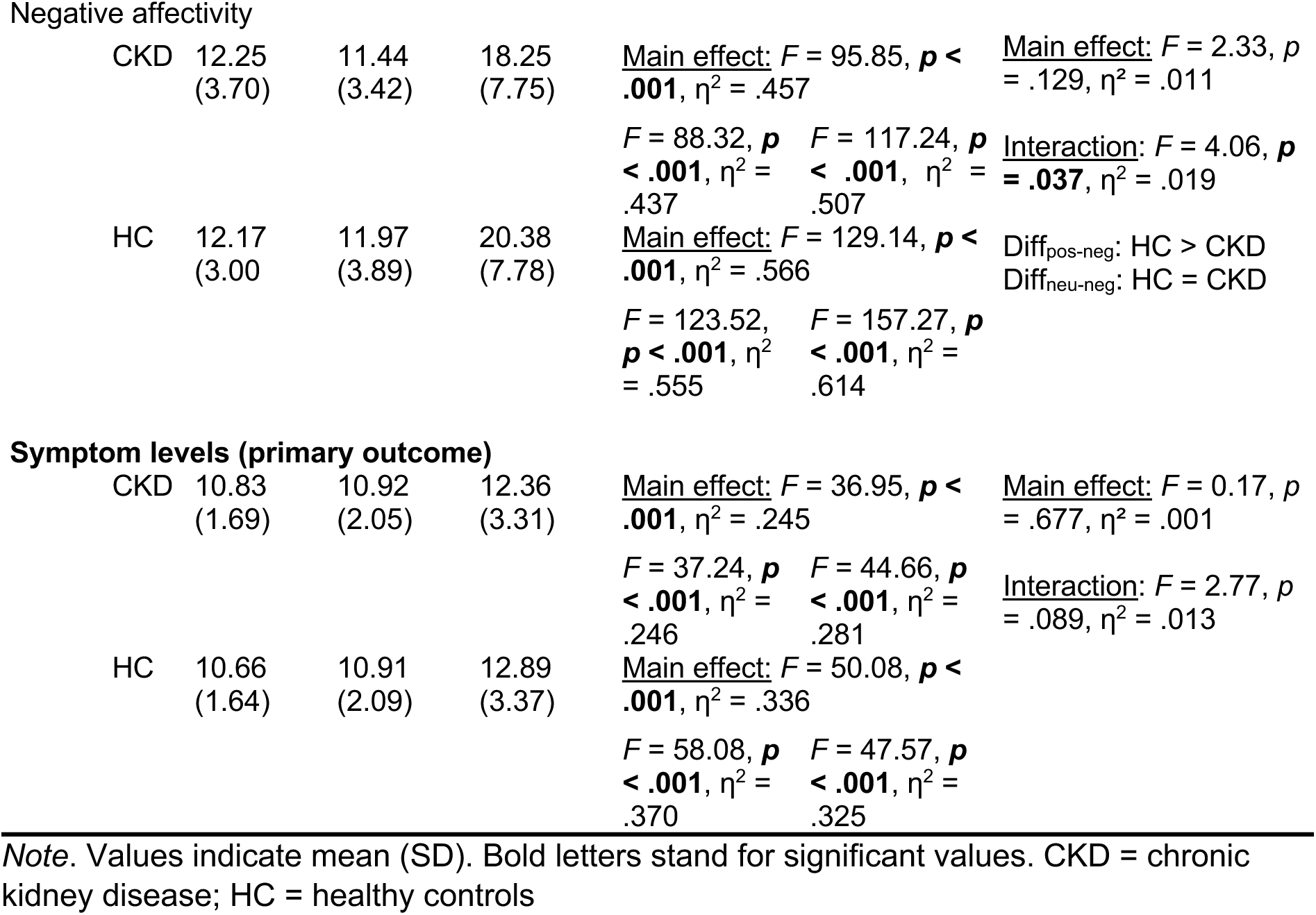
Effects of affect induction on the manipulation check variables and on primary outcome in both groups.

### Symptom levels after negative affect induction (primary outcome)

A significant main effect for picture category emerged, with all participants reporting more symptoms after negative compared to positive and neutral pictures (Table 2, Figure 2). Group affiliation did not moderate symptom levels after affect induction. An independent *t*-test revealed no significant differences in the ASP effect between the CKD (M = 1.43, SD = 2.30) and the HC group ((M = 1.98, SD = 2.87), *t*(213) = −1.54, *p* = .124).

**Figure 2:**
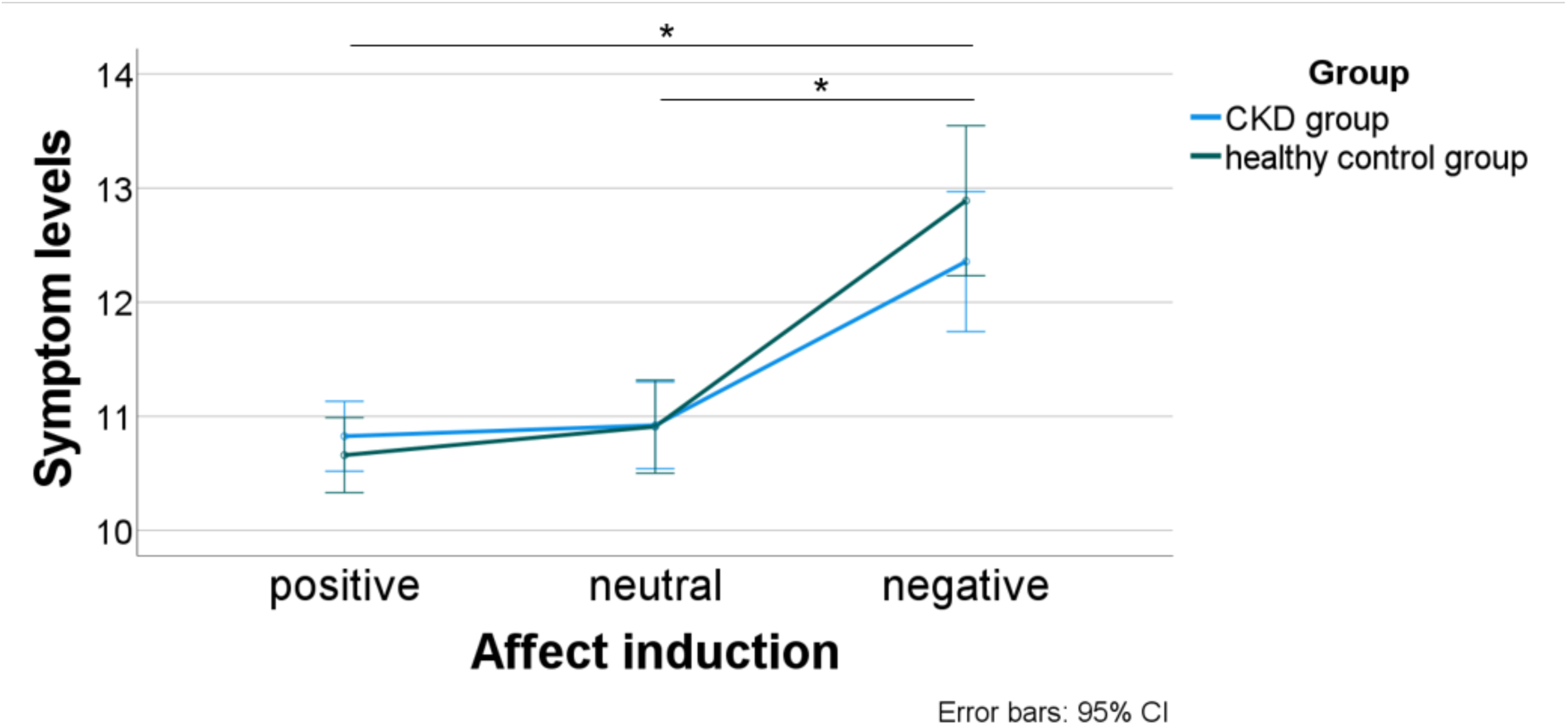
Symptom levels after affect induction. Note. CKD = chronic kidney disease; * p < .05

### Moderators of symptom levels after negative affect induction

As potential moderators, disease severity (CKD group only), habitual symptom burden, trait NA, DIF, emotion regulation and somatosensory amplification were investigated.

*CKD stage* was neither correlated with symptom levels after affect induction (*r* = −.058, *p* = .603), nor did it moderate it (figure 3).

**Figure 3:**
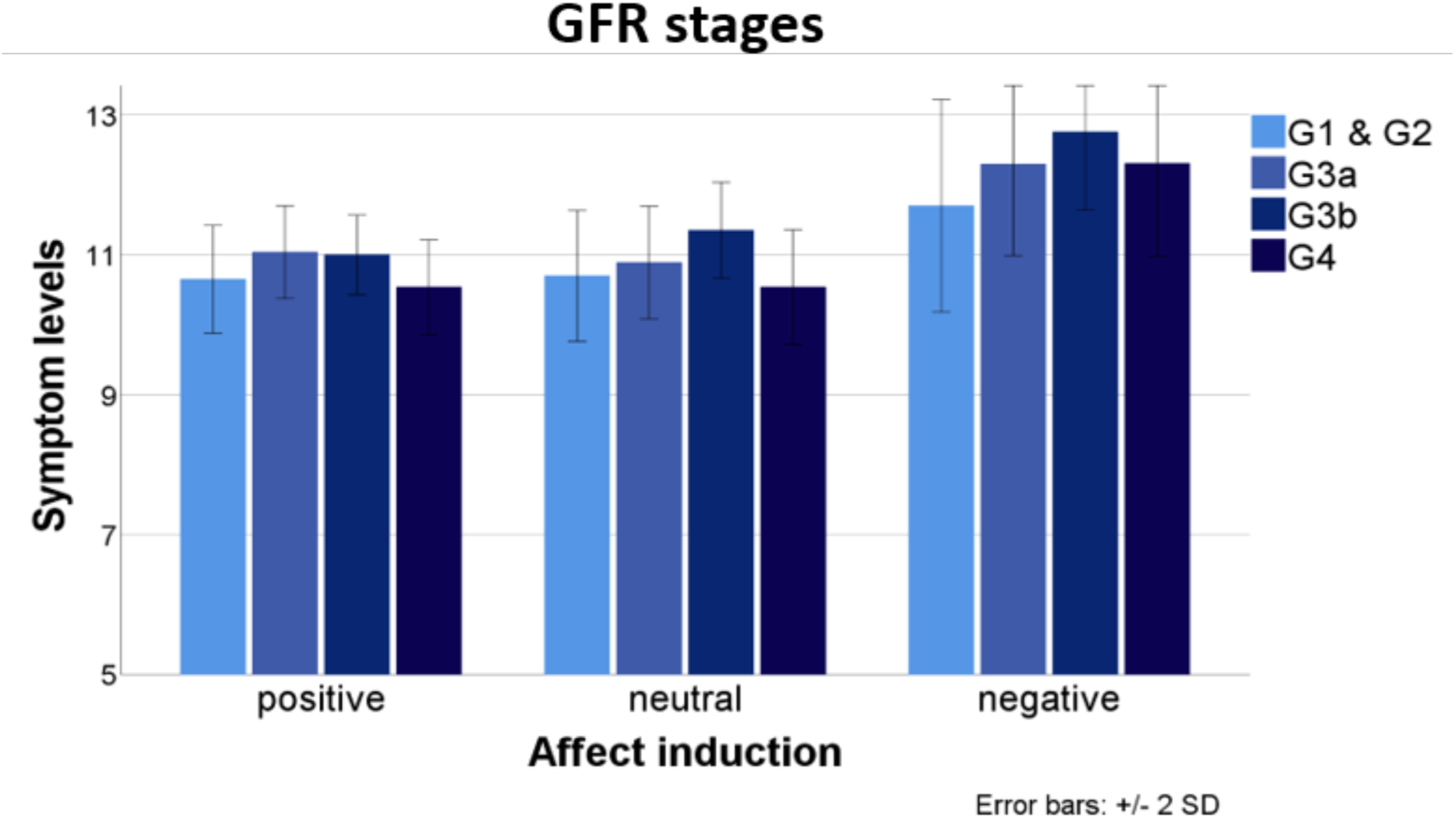
Moderating effect of CKD stage (according to KDIGO guidelines based on GFR [19]) on symptom levels. Note. GFR = glomerular filtration rate; G1-G4 = stages of kidney disease referring to the glomerular filtration rate

Bivariate correlations of symptom levels after affect induction and moderating factors showed that habitual symptom burden (PHQ-15) correlated with symptom levels after negative pictures in both groups (CKD group: *r* = .258, *p* = .005; HC group: *r* = .241, *p* = .016). Higher trait NA correlated with symptom levels (CKD group: *r* = .309, *p* < .001; HC group: *r* = .282, *p* = .004). DIF correlated with symptom levels after negative pictures in both groups (CKD group: *r* = .352, *p* < .001; HC group: *r* = .287, *p* = .004). Somatosensory amplification correlated with symptom levels only in the CKD group (*r* = .276, *p* = .003; HC group: *r* = .014, *p* = .892) (Table S2).

Figure 4 shows the moderator analyses. *High habitual symptom burden* did not moderate symptom levels after negative affect induction. However, a significant main effect showed that individuals with higher habitual symptom burden reported higher symptom levels across all affective picture conditions (Table S3).

**Figure 4:**
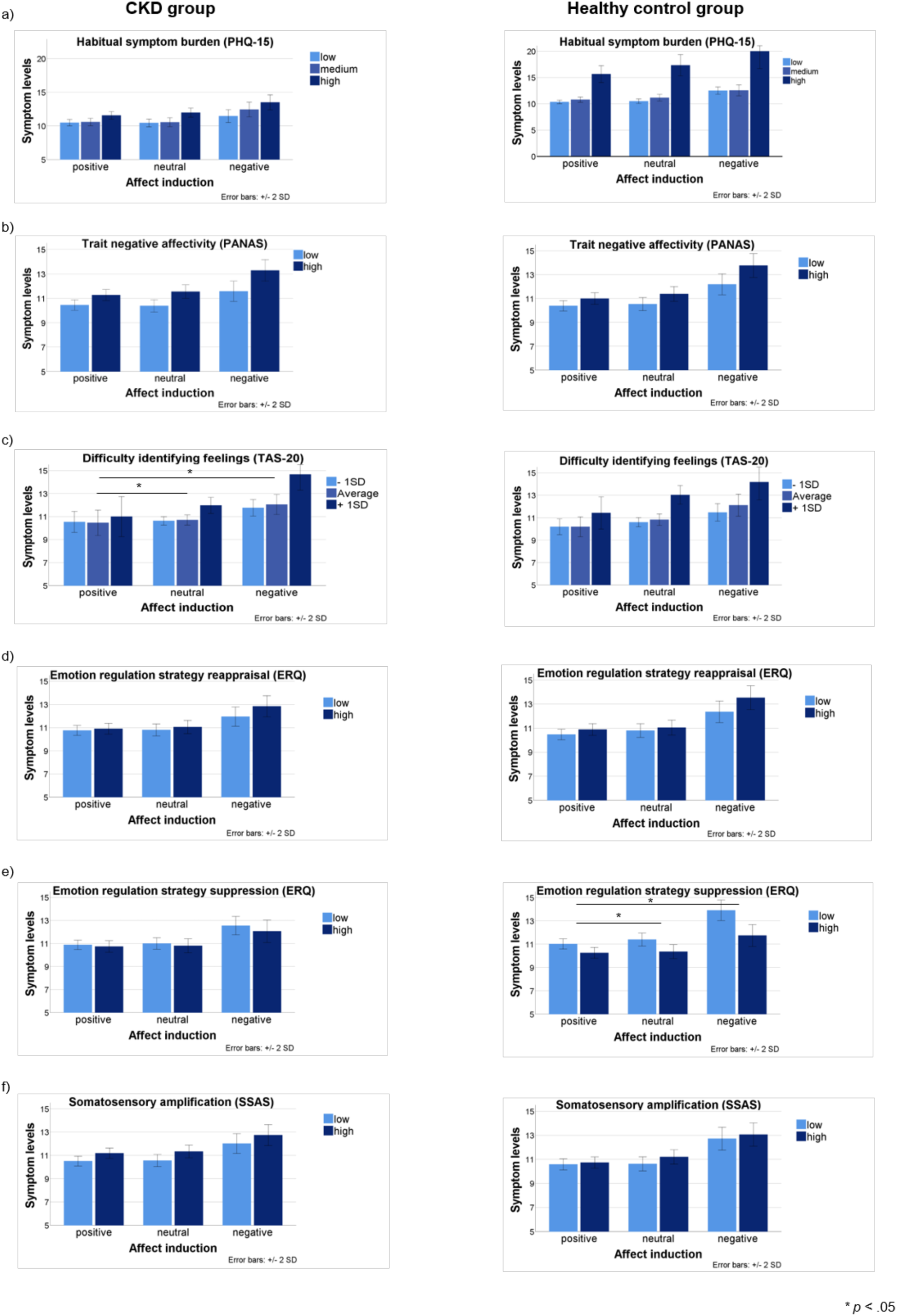
Potential moderators on symptom levels after negative affect induction. Note. F-values refer to the interaction terms. PHQ-15 = Patient Health Questionnaire-15; PANAS = Positive and Negative Affect Schedule; TAS-20 = Toronto Alexithymia Scale 20; ERQ = Emotion Regulation Questionnaire; SSAS = Somatosensory Amplification Scale

*Trait NA* did not significantly moderate symptom levels after negative affect induction, whereas there was a significant main effect in both groups.

*DIF* moderated symptom levels after negative affect induction in the CKD group with higher difficulties leading to increased symptom levels after negative versus positive and neutral pictures and a significant main effect. There was no significant moderation in the HC group but a significant main effect.

*Reappraisal* did not moderate symptom levels. *Suppression* moderated symptom levels only in the HC group with lower symptom levels being associated with more emotional expressive suppression and a significant main effect. Suppression did not moderate symptom levels in the CKD group.

### *Somatosensory amplification* did not moderate symptom levels

Differences in symptom levels by levels of the moderators between groups (CKD & HC) were only found for habitual symptom burden. Post-hoc tests indicated higher symptom levels for participants with high habitual symptom burden in the HC group relative to the CKD group. No Group × Moderator or Group × Affect-induction × Moderator interactions were significant (S3).

Bivariate correlations with the ASP effect only revealed a significant correlation with DIF in the CKD group (*r* = .210, *p* = .025). Other significant correlations such as of suppression and symptom perception after negative pictures disappear (S2, S4).

### Symptom burden at 6 and 12 months in individuals with CKD

In the CKD group, we analysed whether symptom levels after negative affect induction predicted general and CKD-specific symptom burden after 6 and 12 months. Symptom levels correlated with general and CKD-specific symptoms at 6 (general: *r* = .259, *p* = .006, CKD-specific: *r* = .389, *p* < .001) and 12 months (general: *r* = .315, *p* < .001; CKD-specific: r = .219, *p* = .023). The ASP effect correlated with CKD-specific symptom burden at 6 months (*r* = .207, *p* = .03), but not at 12 months. It did not correlate with general symptom burden at 6 and 12 months.

In the prediction model, only baseline habitual symptom burden predicted general symptom burden at 6 and 12 months. CKD-specific symptoms at baseline predicted CKD-specific symptom burden after 6 and 12 months. Both symptom levels after negative affect induction and the ASP effect predicted CKD-specific symptom burden after 6 months (shown for the ASP effect in table 3).

**Table 3:** Associations between the ASP effect and habitual as well as CKD-specific symptom burden after 6 & 12 months in patients with CKD.

| Predictor | Symptom burden at 6 months (PHQ-15) |  | Symptom burden at 12 months (PHQ-15) |  |
| --- | --- | --- | --- | --- |
| n | 110 |  | 108 |  |
|  | Beta | p-values | Beta | p-values |
| | (\|$R^2$\| = .599, $p < .001$, $\Delta R^2 = .607$, $p < .001$) | | (\|$R^2$\| = .593, $p < .001$; $\Delta R^2 = .601$, $p < .001$) | |
| Symptom burden at baseline (PHQ-15) | .773 | <.001 | .772 | <.001 |
| ASP effect | .069 | .258 | .034 | .585 |
|  | CKD-specific symptom burden at 6 months (CKD-SBI) |  | CKD-specific symptom burden at 12 months (CKD-SBI) |  |
| n | 110 |  | 108 |  |
| | (\|$R^2$\| = .658, $p < .001$, $\Delta R^2 = .664$, $p < .001$) | | (\|$R^2$\| = .645, $p < .001$, $\Delta R^2 = .651$, $p < .001$) | |
| CKD-specific symptom burden at baseline (CKD-SBI) | .790 | <.001 | .809 | <.001 |
| ASP effect | .160 | .005 | -.060 | .301 |

Note. PHQ-15 = Patient Health Questionnaire-15; CKD-SBI = Chronic Kidney Disease Symptom Burden Index; ASP = Affect and Symptoms Paradigm

### Exploratory analyses

#### Effect of the ASP on single symptoms

We analysed overall severity of single symptoms (10-item symptom checklist) after positive, neutral and negative pictures using repeated-measures ANOVAs. Seven symptoms were rated significantly higher after negative pictures, two differed between negative and positive pictures. Only muscle pain did not differ between picture categories (Table S5).

## Discussion

This experimental study investigated how symptom perception in patients with CKD can be influenced by inducing negative affect through the ASP compared to healthy controls. As hypothesized, both groups reported higher symptom levels after negative versus positive and neutral pictures. Manipulation checks showed that negative pictures significantly increased NA and arousal, while decreasing valence and feelings of control in both groups. Group affiliation (CKD vs. HC) did not moderate symptom levels after affect induction, indicating a similar response to the ASP in both groups. This highlights the cross-individual consistency of the PPM, which posits that the brain consistently updates predictions minimizing errors and optimizing perception [5]. Thereby, symptom perception can be influenced by affective content even if no somatic input is present. Generally, this aligns with prior research, showing negative affect induction evokes symptoms [9, 13, 14]. To our knowledge, this is the first study investigating the ASP effect in individuals with a chronic symptomatic condition. Based on the PPM, we assumed that the chronic condition could either reduce reactivity to the ASP effect due to elevated symptoms in general and therefore be less salient to additional input or could increase vulnerability to symptom perception through development of strong, precise symptom priors.

Given that the CKD sample responds to negative affect induction with increased symptom levels, one could assume an increased vulnerability to perceiving symptoms through priors. Contradictory to earlier research that found more pronounced symptom reporting in patients with FSS compared to healthy controls, we found increased symptom levels in both groups (CKD and HC). This is in line with recent ASP studies by Petzke et al. [10, 38], who showed the ASP effect in general population samples. The absence of group differences in our sample suggests that chronically ill individuals do not generally respond differently to the ASP. However, our study does not allow causal inferences regarding the effect of somatic input on the ASP effect. Additional research with the ASP in which the somatic input is experimentally manipulated could provide further insight.

Former studies using the ASP investigated younger women (≤ 40 years) [12, 14, 18, 39], whereas our sample was older (CKD group: M = 63 years; HC group: 60 years). Although older adults tend to report more daily symptoms [9, 13, 14], symptom levels after affect induction of our sample are comparable to younger (mostly female) participants [10, 14]. When comparing the CKD group with individuals with FSS in the study of Van den Houte [12], who used the same 10-item symptom checklist, participants with CKD reported less symptoms after each picture category. This suggests that individuals with CKD may experience less intense symptoms after affect induction compared to FSS, suggesting different underlying mechanisms of symptom perception or lower sensitivity to emotional stimuli. As FSS are associated with depressive symptoms, alexithymia and neuroticism [40, 41], symptom perception – as understood within the predictive processing framework – might depend more strongly on priors and therefore lead to more symptoms after affect induction. Differences in age, sex, and comorbidities further distinguish our CKD sample from patients with FSS.

Regarding potential moderators, *disease stage* (CKD group) did not moderate symptom levels after negative affect induction suggesting that kidney function does not influence symptom levels. We showed that affective factors elicit symptoms regardless of the present medical condition, aligning with clinical research showing low associations between kidney function and symptom burden in CKD [21, 23] supporting the disease-overarching concept of PPS [4] Contrary to our hypothesis and recent research [9, 10, 12–14], *high habitual symptom burden* did not moderate symptom levels after affect induction, though a main effect was found in both groups. In healthy controls, only three individuals were classified as high habitual symptom reporters, limiting validity. Likewise, no significant interaction was found between *trait NA* and symptom levels after affect induction, despite correlating bivariately in both groups. A previous study only found an interaction of high-trait-NA/high-habitual-symptom-reporting with symptom levels after affect induction [9]. We used the PHQ-15 measuring habitual symptom burden instead of the Checklist for Symptoms in Daily Life, consisting of 39 symptoms. Possibly no differential effect of high habitual symptom burden in our sample was found because the CKD group might already be burdened by multiple symptoms or older age which may not significantly increase symptoms after negative pictures potentially due to habituation mechanisms according to the PPM [5].

*Difficulties identifying feelings* significantly moderated symptom levels in the CKD group: individuals with greater difficulties identifying feelings reported higher symptom levels after negative pictures, consistent with previous findings [9, 10, 12]. Alexithymia seems to be a risk factor associated with somatic symptom disorder [42] and occurs more frequently in patients on haemodialysis [43] and chronic conditions such as type 2 diabetes [44] or chronic inflammatory bowel disease [45]. It may disrupt interoceptive processing and inaccurate affective predictions, leading to heightened somatic symptom perception in chronic conditions. Individuals with CKD could benefit from treatments training emotion identification reducing prediction errors related to affective and somatic stimuli [12, 46, 47].

HCs who predominantly suppressed their emotions reported less symptoms after negative pictures. *Expressive suppression*, usually associated with lower well-being [48] and reduced emotional adaptability, may have served as short-term protective mechanism, reducing symptom levels after negative stimuli. In contrast, *reappraisal* did not moderate symptom levels but interacted significantly with higher valence in the CKD group highlighting its potential context-dependence of emotion regulation strategies [49–51].

Although *somatosensory amplification* was bivariately linked to symptom levels after negative pictures in the CKD group, it did not moderate it. Since we did not directly add somatosensory input such as pain [52] while watching pictures, we may not have been able to manipulate underlying mechanisms of somatosensory amplification.

Symptom levels after negative affect induction correlated with general and CKD-specific symptoms at 6 and 12 months, suggesting a long-term impact on symptom burden. The extent to which somatic symptoms can be triggered by negative affect seems to be a stable predictor of symptom burden over time, underlining the importance of psychological risk factors in CKD [26] and PSS in general [2]. Thereby, the ASP effect may further indicate the long-term relevance of priors within PPM [53]. However, symptom levels after negative affect induction only predicted CKD-specific symptom burden at 6 months. General and CKD-specific baseline symptoms emerged as strongest predictors of symptom burden at 6 and 12 months. It is premature to recommend the ASP for predicting symptom burden over time, as self-reports on current symptoms provide more reliable data [54]. A recent cross-lagged panel analysis using the ASP at baseline and 18-months follow-up found that habitual symptom burden (PHQ-15) at baseline predicted ASP symptom provocation at follow-up, while ASP symptom provocation at baseline predicted cardiorespiratory symptoms in the PHQ-15 at 18 months [38]. Further research on long-term effects of the ASP effect is needed.

Several limitations must be considered. More precise pre-classification of individuals by habitual symptom burden (low, medium, high) could reinforce symptom levels after negative affect induction. Some participants completed symptom questionnaires (e.g. PHQ-15) immediately before the ASP, which may have triggered unwanted processes such as activating symptom priors that could have interfered with the experimental measurements. As we studied CKD as example symptomatic disease, cross-validation in other chronic diseases as in SOMACROSS [2] may enhance understanding of symptom burden in chronic symptomatic diseases. Using disease-specific symptoms at baseline and after affect induction might capture symptom burden specifically related to the underlying condition. Future research should address these limitations by ensuring precise pre-classification in terms of habitual symptom burden and replicating this experiment in other chronic diseases as in SOMACROSS [2] investigating mechanisms of PSS across diseases.

In conclusion, symptoms in individuals with a chronic somatic condition like CKD responded to the ASP comparably to healthy individuals. Symptom response in the ASP partly predicted symptom burden over time. Notably, while CKD severity did not affect the ASP effect, alexithymia moderated symptom levels in CKD, complementing the biopsychosocial model of symptom perception and offering new directions for improving symptom understanding and treatment.

## Data availability statement

The research data used and analysed during in this study will be shared by the corresponding author upon reasonable request.

## CRediT authorship contribution statement

**Birte Jessen:** conceptualization, methodology, software, formal analysis, investigation, data curation, writing (original draft & review and editing), visualization, project administration. **Meike Shedden-Mora:** conceptualization, methodology, formal analysis, investigation, writing (original draft & review and editing), visualization, supervision, project administration and funding acquisition. **Omer van den Bergh:** conceptualization, methodology, writing (review & editing). **Christian Schmidt-Lauber**: writing (review & editing). **Tobias B. Huber:** writing (review & editing), methodology, funding acquisition. **Bernd Löwe:** writing (review & editing), methodology, funding acquisition. **Michael Witthöft:** writing (review & editing), methodology.

## Funding sources

The study as part of the SOMA.CK study is supported by the German Research Foundation (Deutsche Forschungsgemeinschaft, DFG) (PIs: Meike Shedden-Mora and Tobias B. Huber; project number: 445297796). It is part of the collaborative research unit 5211 (RU 5211) ‘Persistent SOMAtic symptoms ACROSS diseases: from risk factors to modification (SOMACROSS)’ funded by the DFG (spokesperson: BL). For recruiting the healthy control group B.J. received a Mini Grant within the SOMACROSS research unit.

## Submission declaration

The results presented in this article have not been published previously in whole. This article is not under consideration for publication elsewhere. The article’s publication is approved by all authors and tacitly or explicitly by the responsible authorities where the work was carried out.

## Declaration of competing interests

All authors filled in the declaration of interest statements which are attached to this manuscript.

## Declaration of generative AI and AI-assisted technologies in the writing process

During the preparation of this work the authors used DEEPL and ChatGPT in the writing process for better readability. After using this tool, the authors reviewed and edited the content as needed and take full responsibility for the content of the published article.

## Supplementary material

Supplementary material available at…

## Acknowledgements

We are grateful to all patients with CKD and participants in the control group who dedicated their time and energy to support our research. We thank Ines Quent and Simon Dahmlos who contributed to the recruitment of this study, as well as all student research assistants, namely: Efim Lenz, Lisa Autzen, Leah Harding, Verena Fleßner, Ronja Meissner and Niklas Lihl, from the Medical School Hamburg who supported this study. We would also like to thank Sandra Kümmelberg and Fabian Zambiasi for the data management within the SOMACROSS research unit.

## Supplements

S1: Selection of pictures of the IAPS

The following pictures with their specific valence and arousal levels were used:

<u>Positive</u>: 1340, 1463, 1710, 1750, 2058, 2165, 2311, 2340, 2388, 2395, 5001, 5260, 5480, 5700, 5760, 5831, 5833, 7340, 7502, 8502

<u>Neutral</u>: 1121,1560, 1675, 2102, 2214, 2235, 2305, 2396, 2514, 2850, 2880, 5395, 5471, 5520, 5740, 7002, 7004, 7041, 7496, 7640

<u>Negative</u>: 1200, 1932, 2095, 2120, 2692, 2799, 2800, 3500, 5971, 6020, 6313, 6560, 9340, 9342, 9410, 9440, 9561, 9600, 9800, 9911

**Table S1.1.**
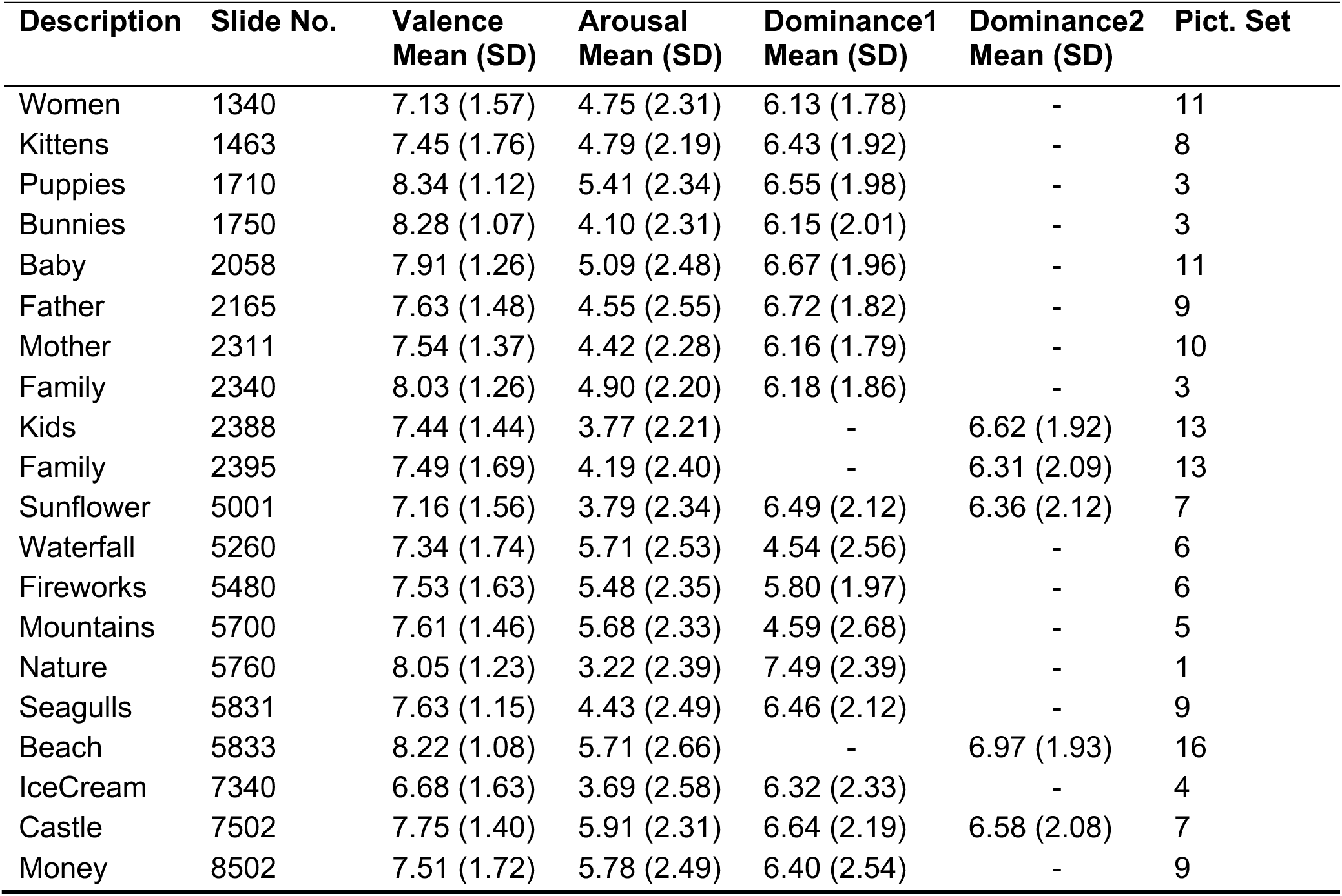
Positive Pictures.

| Description | Slide No. | Valence<br>Mean (SD) | Arousal<br>Mean (SD) | Dominance1<br>Mean (SD) | Dominance2<br>Mean (SD) | Pict. Set |
| --- | --- | --- | --- | --- | --- | --- |
| Women | 1340 | 7.13 (1.57) | 4.75 (2.31) | 6.13 (1.78) | - | 11 |
| Kittens | 1463 | 7.45 (1.76) | 4.79 (2.19) | 6.43 (1.92) | - | 8 |
| Puppies | 1710 | 8.34 (1.12) | 5.41 (2.34) | 6.55 (1.98) | - | 3 |
| Bunnies | 1750 | 8.28 (1.07) | 4.10 (2.31) | 6.15 (2.01) | - | 3 |
| Baby | 2058 | 7.91 (1.26) | 5.09 (2.48) | 6.67 (1.96) | - | 11 |
| Father | 2165 | 7.63 (1.48) | 4.55 (2.55) | 6.72 (1.82) | - | 9 |
| Mother | 2311 | 7.54 (1.37) | 4.42 (2.28) | 6.16 (1.79) | - | 10 |
| Family | 2340 | 8.03 (1.26) | 4.90 (2.20) | 6.18 (1.86) | - | 3 |
| Kids | 2388 | 7.44 (1.44) | 3.77 (2.21) | - | 6.62 (1.92) | 13 |
| Family | 2395 | 7.49 (1.69) | 4.19 (2.40) | - | 6.31 (2.09) | 13 |
| Sunflower | 5001 | 7.16 (1.56) | 3.79 (2.34) | 6.49 (2.12) | 6.36 (2.12) | 7 |
| Waterfall | 5260 | 7.34 (1.74) | 5.71 (2.53) | 4.54 (2.56) | - | 6 |
| Fireworks | 5480 | 7.53 (1.63) | 5.48 (2.35) | 5.80 (1.97) | - | 6 |
| Mountains | 5700 | 7.61 (1.46) | 5.68 (2.33) | 4.59 (2.68) | - | 5 |
| Nature | 5760 | 8.05 (1.23) | 3.22 (2.39) | 7.49 (2.39) | - | 1 |
| Seagulls | 5831 | 7.63 (1.15) | 4.43 (2.49) | 6.46 (2.12) | - | 9 |
| Beach | 5833 | 8.22 (1.08) | 5.71 (2.66) | - | 6.97 (1.93) | 16 |
| IceCream | 7340 | 6.68 (1.63) | 3.69 (2.58) | 6.32 (2.33) | - | 4 |
| Castle | 7502 | 7.75 (1.40) | 5.91 (2.31) | 6.64 (2.19) | 6.58 (2.08) | 7 |
| Money | 8502 | 7.51 (1.72) | 5.78 (2.49) | 6.40 (2.54) | - | 9 |

**Table S1.2.** Neutral Pictures.

| Description | Slide No. | Valence Mean (SD) | Arousal Mean (SD) | Dominance1 Mean (SD) | Dominance2 Mean (SD) | Pict. Set |
| --- | --- | --- | --- | --- | --- | --- |
| Lizard | 1121 | 5.79 (1.61) | 4.83 (1.98) | 5.89 (1.99) | - | 11 |
| Hawk | 1560 | 5.97 (2.32) | 5.51 (2.19) | 5.18 (2.19) | - | 1 |
| Buffalo | 1675 | 5.24 (1.48) | 4.37 (2.15) | - | 4.63 (2.1) | 15 |
| NeuMan | 2102 | 5.16 (0.96) | 3.03 (1.87) | - | 5.8 (1.77) | 16 |
| NeuMan | 2214 | 5.01 (1.12) | 3.46 (1.97) | 5.98 (1.69) | - | 11 |
| Butcher | 2235 | 5.64 (1.27) | 3.36 (1.92) | - | 5.83 (1.91) | 13 |
| Woman | 2305 | 5.41 (1.12) | 3.63 (2.04) | - | 5.53 (1.49) | 16 |
| Couple | 2396 | 4.91 (1.05) | 3.34 (1.83) | - | 5.59 (1.59) | 16 |
| Woman | 2514 | 5.19 (1.09) | 3.50 (1.81) | 5.85 (1.83) | - | 11 |
| Tourist | 2850 | 5.22 (1.39) | 3.00 (1.94) | 5.87 (1.97) | - | 10 |
| Shadow | 2880 | 5.18 (1.44) | 2.96 (1.94) | 6.01 (2.05) | - | 9 |
| Boat | 5395 | 5.34 (1.21) | 4.23 (2.03) | 5.23 (2.01) | - | 12 |
| Satellite | 5471 | 5.21 (1.18) | 3.26 (2.05) | - | 5.63 (1.82) | 16 |
| Mushroom | 5520 | 5.33 (1.49) | 2.95 (2.42) | 6.43 (2.42) | - | 1 |
| Plant | 5740 | 5.21 (1.38) | 2.59 (1.99) | 6.27 (2.21) | - | 10 |
| Towel | 7002 | 4.97 (0.97) | 3.16 (2.00) | 6.25 (1.93) | 6.48 (1.94) | 7 |
| Spoon | 7004 | 5.04 (0.60) | 2.00 (1.66) | 6.74 (1.99) | - | 9 |
| Baskets | 7041 | 4.99 (1.12) | 2.60 (1.78) | - | 6.35 (2.14) | 14 |
| Street | 7496 | 5.92 (1.66) | 4.84 (1.99) | 5.55 (1.72) | - | 12 |
| Skyscraper | 7640 | 5.00 (1.31) | 6.03 (2.46) | 3.82 (2.27) | - | 6 |

**Table S1.3.**
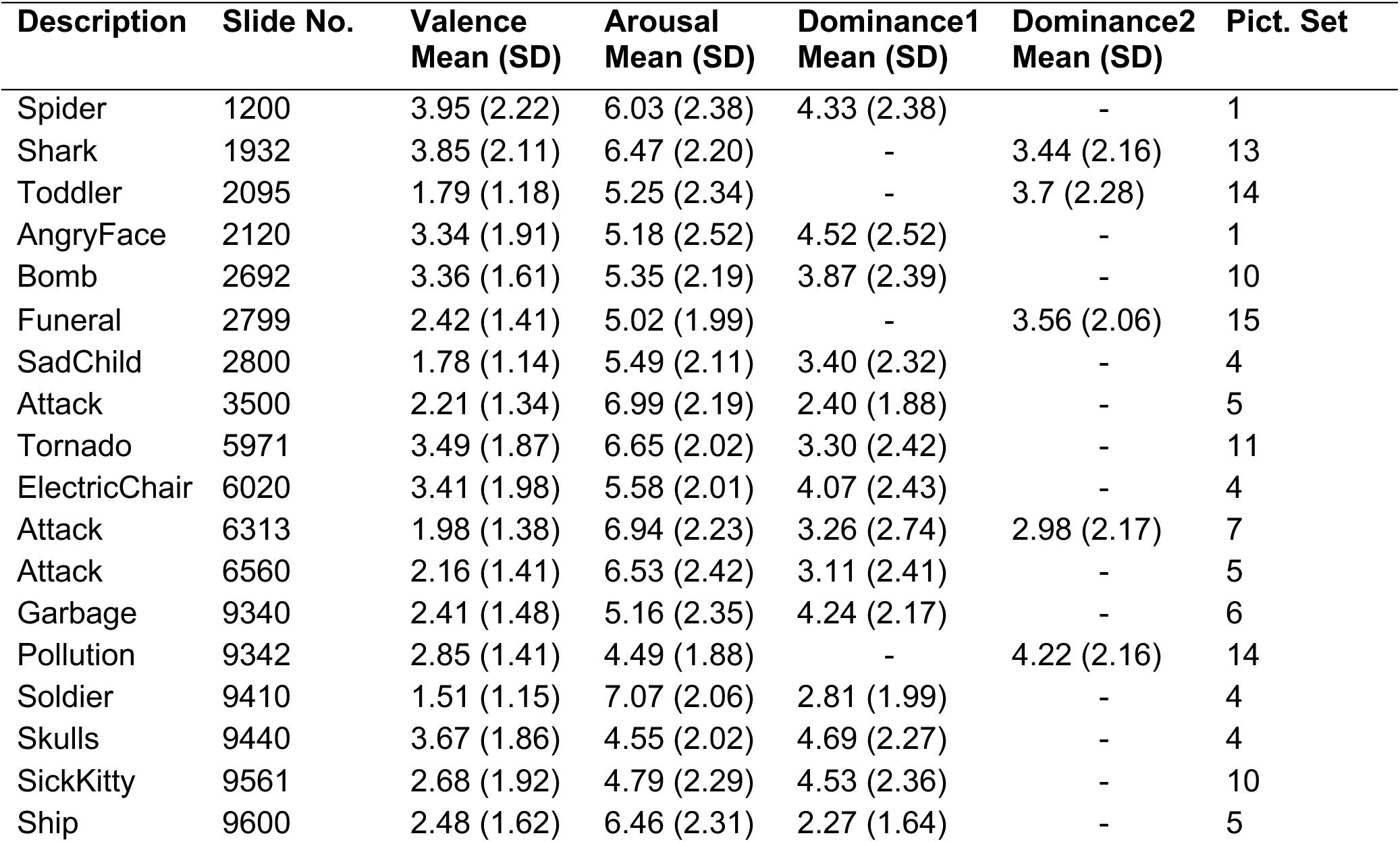
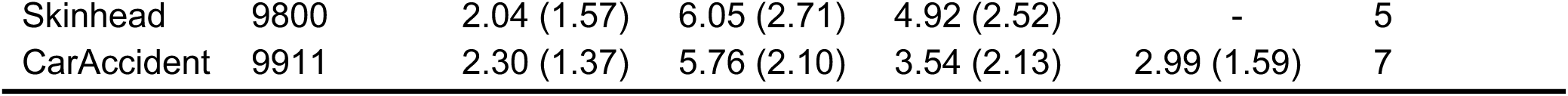
Negative Pictures.

| Description | Slide No. | Valence Mean (SD) | Arousal Mean (SD) | Dominance1 Mean (SD) | Dominance2 Mean (SD) | Pict. Set |
| --- | --- | --- | --- | --- | --- | --- |
| Spider | 1200 | 3.95 (2.22) | 6.03 (2.38) | 4.33 (2.38) | - | 1 |
| Shark | 1932 | 3.85 (2.11) | 6.47 (2.20) | - | 3.44 (2.16) | 13 |
| Toddler | 2095 | 1.79 (1.18) | 5.25 (2.34) | - | 3.7 (2.28) | 14 |
| AngryFace | 2120 | 3.34 (1.91) | 5.18 (2.52) | 4.52 (2.52) | - | 1 |
| Bomb | 2692 | 3.36 (1.61) | 5.35 (2.19) | 3.87 (2.39) | - | 10 |
| Funeral | 2799 | 2.42 (1.41) | 5.02 (1.99) | - | 3.56 (2.06) | 15 |
| SadChild | 2800 | 1.78 (1.14) | 5.49 (2.11) | 3.40 (2.32) | - | 4 |
| Attack | 3500 | 2.21 (1.34) | 6.99 (2.19) | 2.40 (1.88) | - | 5 |
| Tornado | 5971 | 3.49 (1.87) | 6.65 (2.02) | 3.30 (2.42) | - | 11 |
| ElectricChair | 6020 | 3.41 (1.98) | 5.58 (2.01) | 4.07 (2.43) | - | 4 |
| Attack | 6313 | 1.98 (1.38) | 6.94 (2.23) | 3.26 (2.74) | 2.98 (2.17) | 7 |
| Attack | 6560 | 2.16 (1.41) | 6.53 (2.42) | 3.11 (2.41) | - | 5 |
| Garbage | 9340 | 2.41 (1.48) | 5.16 (2.35) | 4.24 (2.17) | - | 6 |
| Pollution | 9342 | 2.85 (1.41) | 4.49 (1.88) | - | 4.22 (2.16) | 14 |
| Soldier | 9410 | 1.51 (1.15) | 7.07 (2.06) | 2.81 (1.99) | - | 4 |
| Skulls | 9440 | 3.67 (1.86) | 4.55 (2.02) | 4.69 (2.27) | - | 4 |
| SickKitty | 9561 | 2.68 (1.92) | 4.79 (2.29) | 4.53 (2.36) | - | 10 |
| Ship | 9600 | 2.48 (1.62) | 6.46 (2.31) | 2.27 (1.64) | - | 5 |
| Skinhead | 9800 | 2.04 (1.57) | 6.05 (2.71) | 4.92 (2.52) | - | 5 |
| CarAccident | 9911 | 2.30 (1.37) | 5.76 (2.10) | 3.54 (2.13) | 2.99 (1.59) | 7 |

**Table S2:**
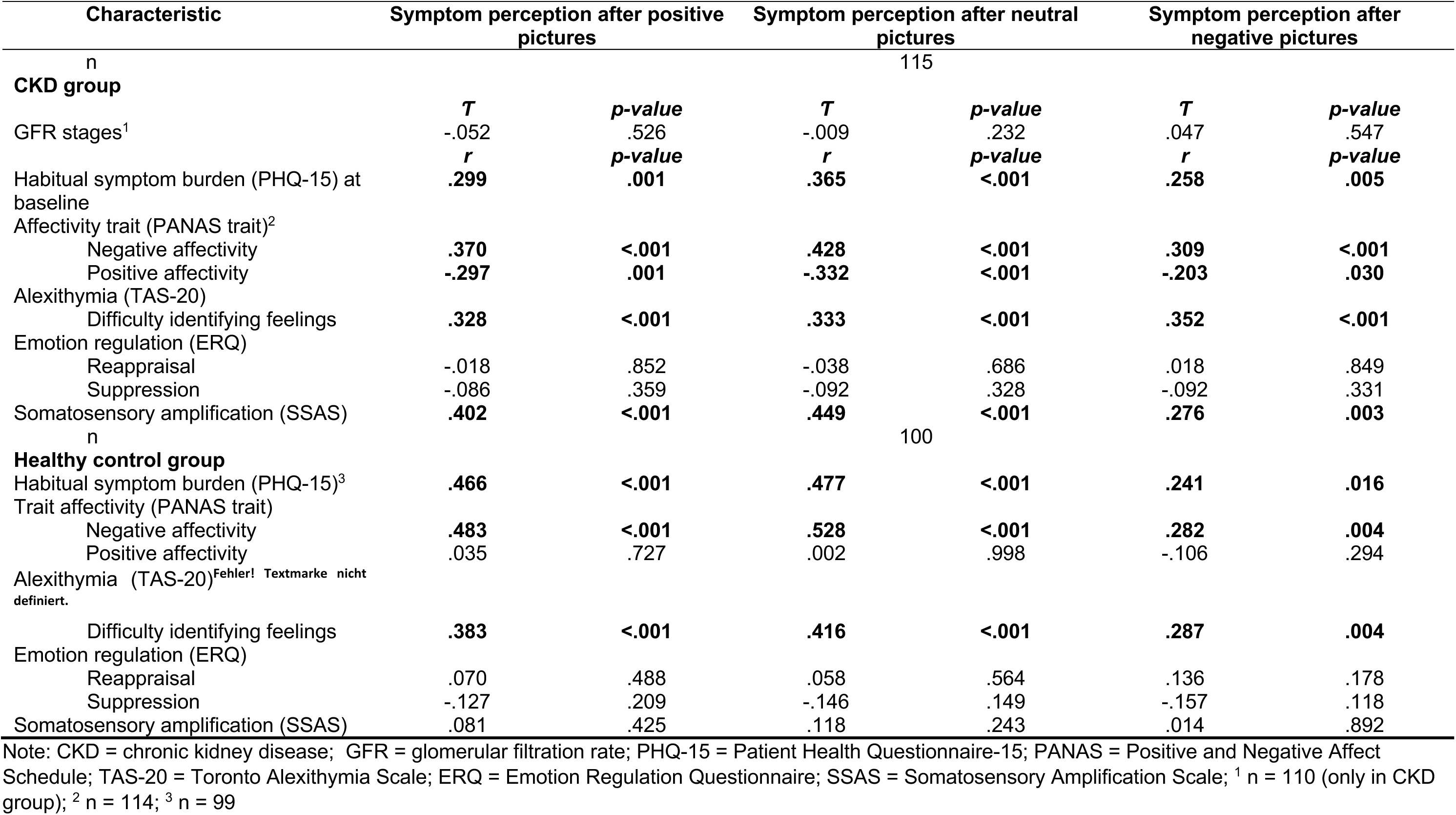
Correlations of symptom levels after affect induction and moderating factors per group.

**Table S3:**
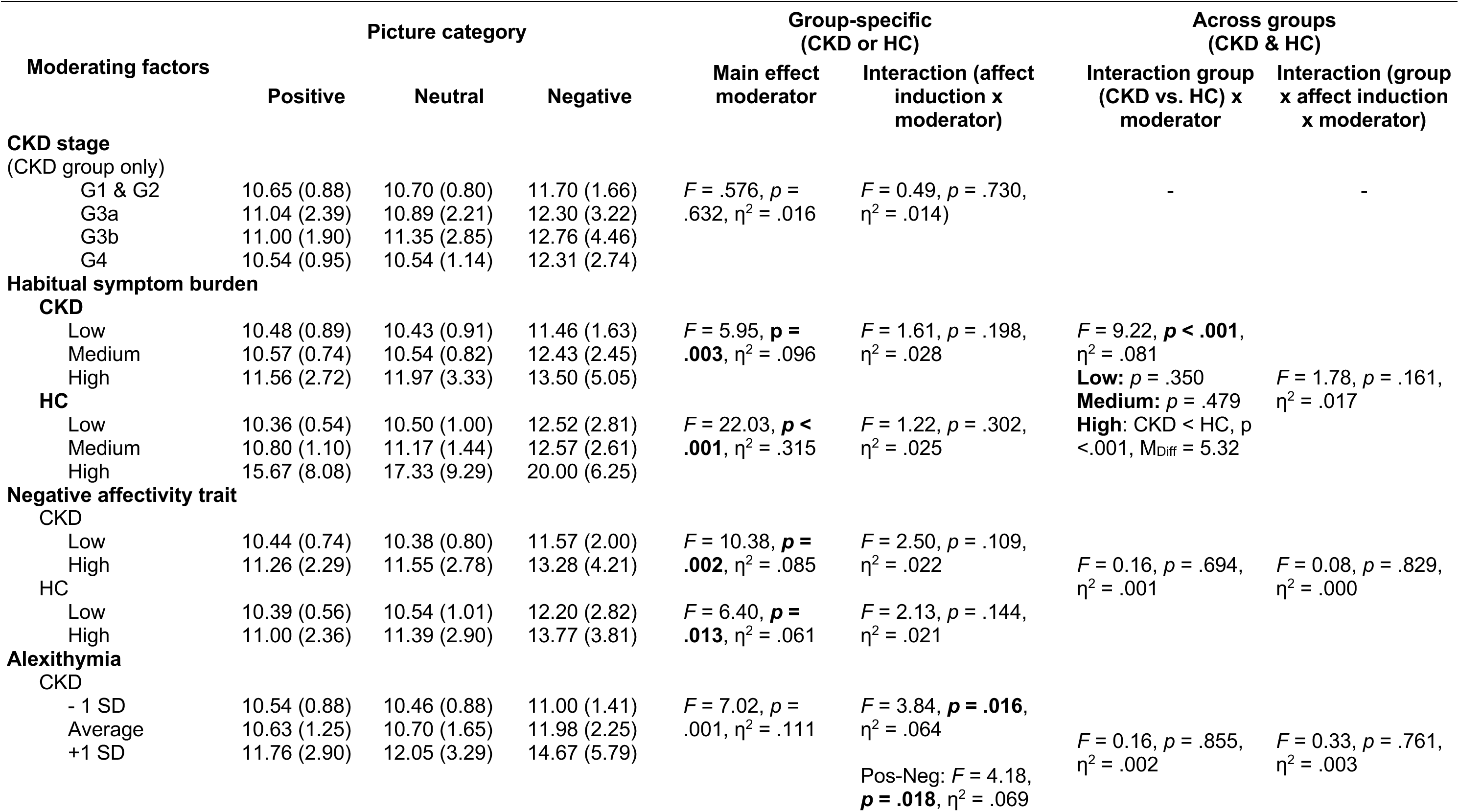
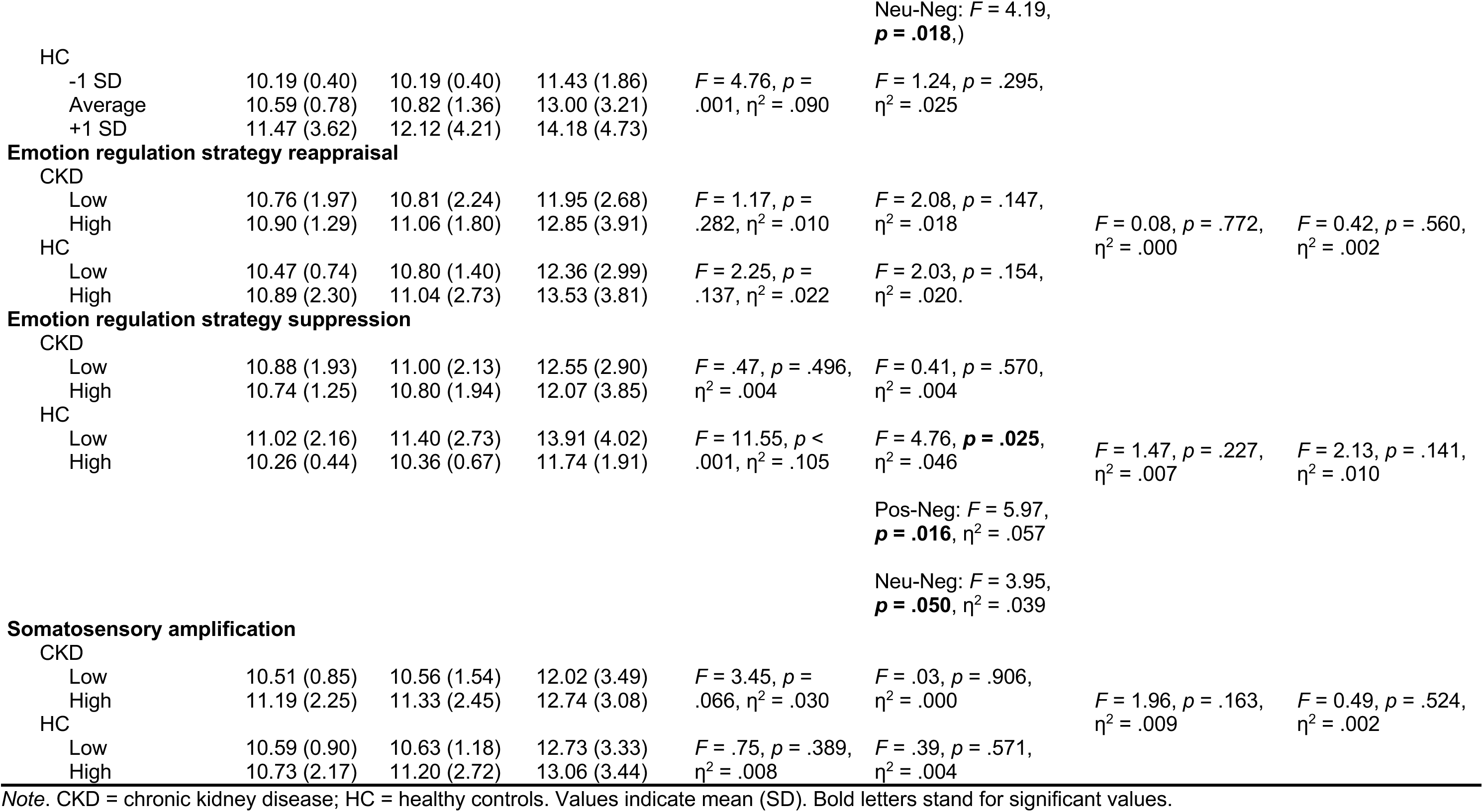
Group-specific and cross-group comparison (CKD vs. HC) regarding main differences in moderators and their interaction with picture category.

| Moderating factors | Picture category |  |  | Group-specific<br>(CKD or HC) |  | Across groups<br>(CKD & HC) |  |
| --- | --- | --- | --- | --- | --- | --- | --- |
|  | Positive | Neutral | Negative | Main effect<br>moderator | Interaction (affect<br>induction x<br>moderator) | Interaction group<br>(CKD vs. HC) x<br>moderator | Interaction (group<br>x affect induction<br>x moderator) |
| **CKD stage**<br>(CKD group only) |  |  |  |  |  |  |  |
| G1 & G2 | 10.65 (0.88) | 10.70 (0.80) | 11.70 (1.66) | $F = .576, p = .632, \eta^2 = .016$ | $F = 0.49, p = .730, \eta^2 = .014$ | - | - |
| G3a | 11.04 (2.39) | 10.89 (2.21) | 12.30 (3.22) |  |  |  |  |
| G3b | 11.00 (1.90) | 11.35 (2.85) | 12.76 (4.46) |  |  |  |  |
| G4 | 10.54 (0.95) | 10.54 (1.14) | 12.31 (2.74) |  |  |  |  |
| **Habitual symptom burden** |  |  |  |  |  |  |  |
| **CKD** |  |  |  |  |  |  |  |
| Low | 10.48 (0.89) | 10.43 (0.91) | 11.46 (1.63) | $F = 5.95, p = .003, \eta^2 = .096$ | $F = 1.61, p = .198, \eta^2 = .028$ | $F = 9.22, p < .001, \eta^2 = .081$<br>**Low:** $p = .350$<br>**Medium:** $p = .479$<br>**High:** CKD < HC, $p < .001, M_{Diff} = 5.32$ | $F = 1.78, p = .161, \eta^2 = .017$ |
| Medium | 10.57 (0.74) | 10.54 (0.82) | 12.43 (2.45) |  |  |  |  |
| High | 11.56 (2.72) | 11.97 (3.33) | 13.50 (5.05) |  |  |  |  |
| **HC** |  |  |  |  |  |  |  |
| Low | 10.36 (0.54) | 10.50 (1.00) | 12.52 (2.81) | $F = 22.03, p < .001, \eta^2 = .315$ | $F = 1.22, p = .302, \eta^2 = .025$ | | |
| Medium | 10.80 (1.10) | 11.17 (1.44) | 12.57 (2.61) |  |  |  |  |
| High | 15.67 (8.08) | 17.33 (9.29) | 20.00 (6.25) |  |  |  |  |
| **Negative affectivity trait** |  |  |  |  |  |  |  |
| **CKD** |  |  |  |  |  |  |  |
| Low | 10.44 (0.74) | 10.38 (0.80) | 11.57 (2.00) | $F = 10.38, p = .002, \eta^2 = .085$ | $F = 2.50, p = .109, \eta^2 = .022$ | $F = 0.16, p = .694, \eta^2 = .001$ | $F = 0.08, p = .829, \eta^2 = .000$ |
| High | 11.26 (2.29) | 11.55 (2.78) | 13.28 (4.21) |  |  |  |  |
| **HC** |  |  |  |  |  |  |  |
| Low | 10.39 (0.56) | 10.54 (1.01) | 12.20 (2.82) | $F = 6.40, p = .013, \eta^2 = .061$ | $F = 2.13, p = .144, \eta^2 = .021$ | | |
| High | 11.00 (2.36) | 11.39 (2.90) | 13.77 (3.81) |  |  |  |  |
| **Alexithymia** |  |  |  |  |  |  |  |
| **CKD** |  |  |  |  |  |  |  |
| - 1 SD | 10.54 (0.88) | 10.46 (0.88) | 11.00 (1.41) | $F = 7.02, p = .001, \eta^2 = .111$ | $F = 3.84, p = .016, \eta^2 = .064$<br>Pos-Neg: $F = 4.18, p = .018, \eta^2 = .069$ | $F = 0.16, p = .855, \eta^2 = .002$ | $F = 0.33, p = .761, \eta^2 = .003$ |
| Average | 10.63 (1.25) | 10.70 (1.65) | 11.98 (2.25) |  |  |  |  |
| +1 SD | 11.76 (2.90) | 12.05 (3.29) | 14.67 (5.79) |  |  |  |  |

|  |  |  |  |  |  |  |  |  |
| --- | --- | --- | --- | --- | --- | --- | --- | --- |
| | | | | | Neu-Neg: $F = 4.19$,<br>$p = .018$,) | | | |
| HC |  |  |  |  |  |  |  |  |
| -1 SD | 10.19 (0.40) | 10.19 (0.40) | 11.43 (1.86) | $F = 4.76$, $p =$ | $F = 1.24$, $p = .295$, | | | |
| Average | 10.59 (0.78) | 10.82 (1.36) | 13.00 (3.21) | .001, $\eta^2 = .090$ | $\eta^2 = .025$ | | | |
| +1 SD | 11.47 (3.62) | 12.12 (4.21) | 14.18 (4.73) |  |  |  |  |  |
| Emotion regulation strategy reappraisal |  |  |  |  |  |  |  |  |
| CKD |  |  |  |  |  |  |  |  |
| Low | 10.76 (1.97) | 10.81 (2.24) | 11.95 (2.68) | $F = 1.17$, $p =$ | $F = 2.08$, $p = .147$, | | | |
| High | 10.90 (1.29) | 11.06 (1.80) | 12.85 (3.91) | .282, $\eta^2 = .010$ | $\eta^2 = .018$ | $F = 0.08$, $p = .772$, | $F = 0.42$, $p = .560$, | |
| | | | | | | $\eta^2 = .000$ | $\eta^2 = .002$ | |
| HC |  |  |  |  |  |  |  |  |
| Low | 10.47 (0.74) | 10.80 (1.40) | 12.36 (2.99) | $F = 2.25$, $p =$ | $F = 2.03$, $p = .154$, | | | |
| High | 10.89 (2.30) | 11.04 (2.73) | 13.53 (3.81) | .137, $\eta^2 = .022$ | $\eta^2 = .020$. | | | |
| Emotion regulation strategy suppression |  |  |  |  |  |  |  |  |
| CKD |  |  |  |  |  |  |  |  |
| Low | 10.88 (1.93) | 11.00 (2.13) | 12.55 (2.90) | $F = .47$, $p = .496$, | $F = 0.41$, $p = .570$, | | | |
| High | 10.74 (1.25) | 10.80 (1.94) | 12.07 (3.85) | $\eta^2 = .004$ | $\eta^2 = .004$ | | | |
| HC |  |  |  |  |  |  |  |  |
| Low | 11.02 (2.16) | 11.40 (2.73) | 13.91 (4.02) | $F = 11.55$, $p <$ | $F = 4.76$, $p = .025$, | $F = 1.47$, $p = .227$, | $F = 2.13$, $p = .141$, | |
| High | 10.26 (0.44) | 10.36 (0.67) | 11.74 (1.91) | .001, $\eta^2 = .105$ | $\eta^2 = .046$ | $\eta^2 = .007$ | $\eta^2 = .010$ | |
| | | | | | Pos-Neg: $F = 5.97$,<br>$p = .016$, $\eta^2 = .057$ | | | |
| | | | | | Neu-Neg: $F = 3.95$,<br>$p = .050$, $\eta^2 = .039$ | | | |
| Somatosensory amplification |  |  |  |  |  |  |  |  |
| CKD |  |  |  |  |  |  |  |  |
| Low | 10.51 (0.85) | 10.56 (1.54) | 12.02 (3.49) | $F = 3.45$, $p =$ | $F = .03$, $p = .906$, | | | |
| High | 11.19 (2.25) | 11.33 (2.45) | 12.74 (3.08) | .066, $\eta^2 = .030$ | $\eta^2 = .000$ | $F = 1.96$, $p = .163$, | $F = 0.49$, $p = .524$, | |
| | | | | | | $\eta^2 = .009$ | $\eta^2 = .002$ | |
| HC |  |  |  |  |  |  |  |  |
| Low | 10.59 (0.90) | 10.63 (1.18) | 12.73 (3.33) | $F = .75$, $p = .389$, | $F = .39$, $p = .571$, | | | |
| High | 10.73 (2.17) | 11.20 (2.72) | 13.06 (3.44) | $\eta^2 = .008$ | $\eta^2 = .004$ | | | |

Note. CKD = chronic kidney disease; HC = healthy controls. Values indicate mean (SD). Bold letters stand for significant values.

**Table S4:** Correlations of moderating factors with the ASP effect.

| Characteristic | Overall |  | CKD group |  | Healthy control group |  |
| --- | --- | --- | --- | --- | --- | --- |
|  |  |  | ASP effect |  |  |  |
| n | 215 |  | 115 |  | 100 |  |
|  | *r/T* | *p-value* | *r/T* | *p-value* | *r/T* | *p-value* |
| **Moderating factors** |  |  |  |  |  |  |
| GFR stages<sup>1</sup> | - | - | .056 | .474 | - | - |
| Habitual symptom burden (PHQ-15) at baseline | -.035 | .612 | .046 | .626 | -.098 | .333 |
| Affectivity trait (PANAS trait)<sup>2</sup> |  |  |  |  |  |  |
| Positive affectivity | -.024 | .727 | .004 | .964 | -.125 | .214 |
| Negative affectivity | .007 | .924 | .063 | .505 | -.054 | .594 |
| Alexithymia (TAS-20)<sup>3</sup> |  |  |  |  |  |  |
| Difficulty identifying feelings | .107 | .118 | **.210** | **.025** | .032 | .750 |
| Emotion regulation (ERQ) |  |  |  |  |  |  |
| Reappraisal | .098 | .150 | .060 | .526 | .117 | .247 |
| Suppression | -.086 | .208 | -.050 | .598 | -.078 | .438 |
| Somatosensory amplification (SSAS) | -.004 | .952 | -.004 | .969 | -.070 | .490 |

Note: ASP = Affect and Symptoms Paradigm; CKD = chronic kidney disease; GFR = glomerular filtration rate; PHQ-15 = Patient Health Questionnaire-15; PANAS = Positive and Negative Affect Schedule; TAS-20 = Toronto Alexithymia Scale; ERQ = Emotion Regulation Questionnaire; SSAS = Somatosensory Amplification Scale; <sup>1</sup> n = 110 (only in CKD group); <sup>2</sup> n = 114; <sup>3</sup> n = 99

**Table S5:** Overall number of symptoms of the 10-item symptom checklist.

| Characteristic | Positive picture category | Neutral picture category | Negative picture category | Repeated measures ANOVA | Diff<sub>pos-neg</sub> [95%-CI] | Diff<sub>neu-neg</sub> [95%-CI] |
| --- | --- | --- | --- | --- | --- | --- |
| N |  |  |  | 215 |  |  |
| **Single symptoms** |  |  |  |  |  |  |
| Tightness in the chest | 1.06 ± 0.33 | 1.06 ± 0.25 | 1.51 ± 0.69 | $F(1.38, 21.52) = 79.44$,<br>$p < .001$, $\eta^2 = .271$ | -.46, $p < .001$,<br>[-.58, -.33] | -.46, $p < .001$,<br>[-.56, -.35] |
| Heart pounding | 1.24 ± 0.46 | 1.14 ± 0.36 | 1.66 ± 0.70 | $F(1.66, 19.82) = 86.37$,<br>$p < .001$, $\eta^2 = .288$ | -.42, $p < .001$,<br>[-.54, -.31] | -.52, $p < .001$,<br>[-.63, -.41] |
| Stomach pain | 1.02 ± 0.14 | 1.03 ± 0.19 | 1.18 ± 0.46 | $F(1.22, 2.78) = 24.20$,<br>$p < .001$, $\eta^2 = .102$ | -.16, $p < .001$,<br>[-.23, -.09] | -.15, $p < .001$,<br>[-.22, -.08] |
| Headaches | 1.04 ± 0.30 | 1.08 ± 0.41 | 1.15 ± 0.46 | $F(1.62, 0.84) = 10.99$,<br>$p < .001$, $\eta^2 = .049$ | -.11, $p < .001$,<br>[-.18, -.04] | -.07, $p = .005$,<br>[-.12, -.02] |
| Fatigue | 1.23 ± 0.57 | 1.38 ± 0.69 | 1.37 ± 0.62 | $F(1.87, 1.60) = 10.84$,<br>$p < .001$, $\eta^2 = .048$ | -.14, $p < .001$,<br>[-.22, -.06] | .01, $p = 1.00$,<br>[-.09, .11] |
| Shortness of breath | 1.05 ± 0.32 | 1.07 ± 0.36 | 1.18 ± 0.50 | $F(1.33, 1.72) = 18.19$,<br>$p < .001$, $\eta^2 = .078$ | -.14, $p < .001$,<br>[-.20, -.07] | -.12, $p < .001$,<br>[-.18, -.05] |
| Heart racing | 1.04 ± 0.19 | 1.04 ± 0.20 | 1.22 ± 0.48 | $F(1.36, 3.55) = 30.62$,<br>$p < .001$, $\eta^2 = .125$ | -.19, $p < .001$,<br>[-.26, -.11] | -.18, $p < .001$,<br>[-.25, -.11] |
| Nausea | 1.00 ± 0.00 | 1.00 ± 0.68 | 1.17 ± 0.48 | $F(1.03, 3.80) = 25.04$,<br>$p < .001$, $\eta^2 = .105$ | -.17, $p < .001$,<br>[-.25, -.09] | -.16, $p < .001$,<br>[-.24, -.08] |
| Dizziness | 1.02 ± 0.18 | 1.04 ± 0.24 | 1.07 ± 0.33 | $F(1.57, 0.18) = 4.99$,<br>$p = .013$, $\eta^2 = .023$ | -.05, $p = .022$,<br>[-.10, -.01] | -.03, $p = .212$,<br>[-.08, .01] |
| Muscle pain | 1.06 ± 0.34 | 1.08 ± 0.37 | 1.09 ± 0.36 | $F(1.88, 0.46) = 1.76$,<br>$p = .176$, $\eta^2 = .008$ | -.03, $p = .250$,<br>[-.07, .01] | -.01, $p = 1.00$,<br>[-.05, .03] |

2 Note. Values indicate mean ± SD.

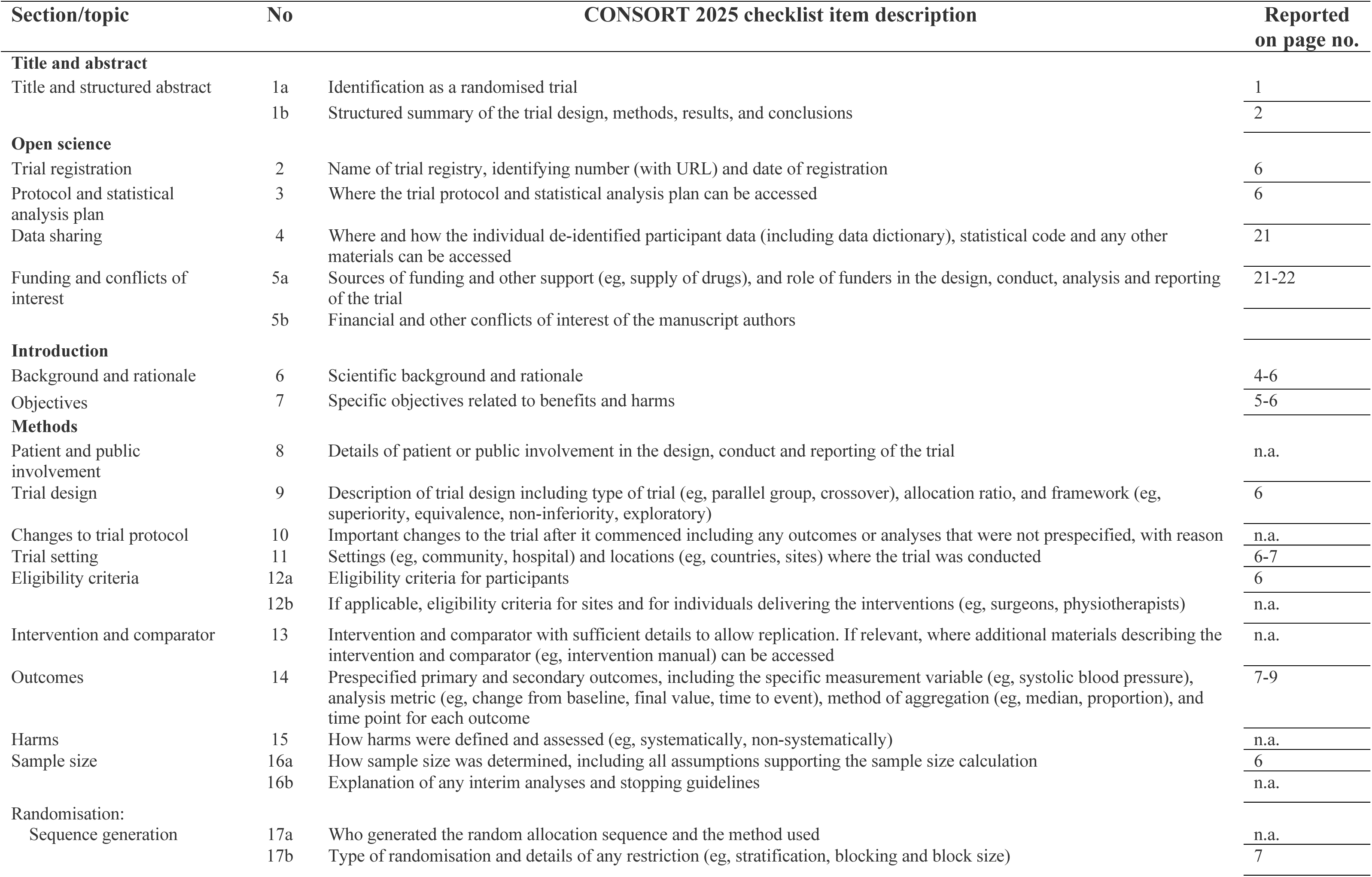
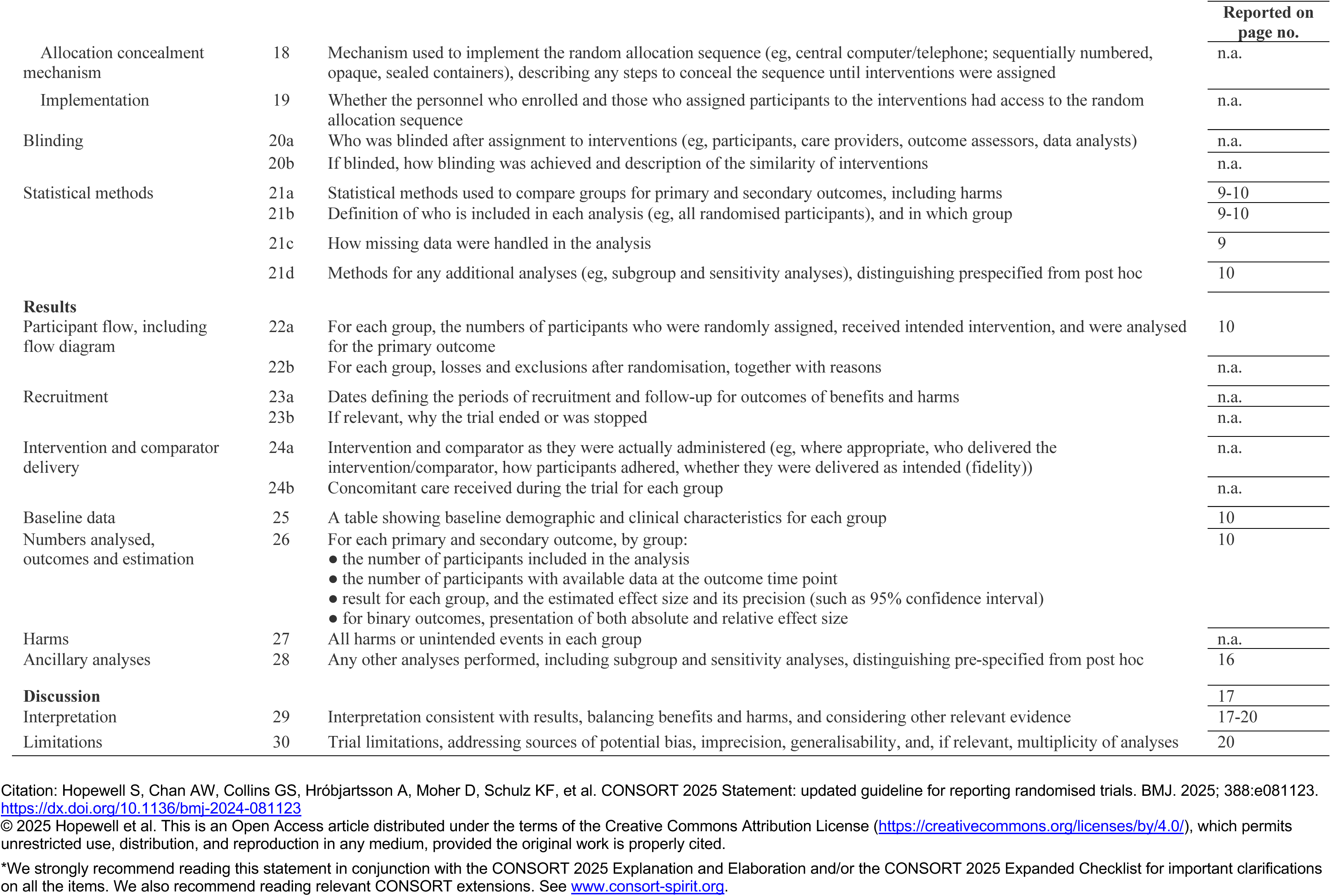

2 Note. Values indicate mean ± SD.
| Section/topic | No | CONSORT 2025 checklist item description | Reported on page no. |
| --- | --- | --- | --- |
| **Title and abstract** |  |  |  |
| Title and structured abstract | 1a | Identification as a randomised trial | 1 |
|  | 1b | Structured summary of the trial design, methods, results, and conclusions | 2 |
| **Open science** |  |  |  |
| Trial registration | 2 | Name of trial registry, identifying number (with URL) and date of registration | 6 |
| Protocol and statistical analysis plan | 3 | Where the trial protocol and statistical analysis plan can be accessed | 6 |
| Data sharing | 4 | Where and how the individual de-identified participant data (including data dictionary), statistical code and any other materials can be accessed | 21 |
| Funding and conflicts of interest | 5a | Sources of funding and other support (eg, supply of drugs), and role of funders in the design, conduct, analysis and reporting of the trial | 21-22 |
|  | 5b | Financial and other conflicts of interest of the manuscript authors |  |
| **Introduction** |  |  |  |
| Background and rationale | 6 | Scientific background and rationale | 4-6 |
| Objectives | 7 | Specific objectives related to benefits and harms | 5-6 |
| **Methods** |  |  |  |
| Patient and public involvement | 8 | Details of patient or public involvement in the design, conduct and reporting of the trial | n.a. |
| Trial design | 9 | Description of trial design including type of trial (eg, parallel group, crossover), allocation ratio, and framework (eg, superiority, equivalence, non-inferiority, exploratory) | 6 |
| Changes to trial protocol | 10 | Important changes to the trial after it commenced including any outcomes or analyses that were not prespecified, with reason | n.a. |
| Trial setting | 11 | Settings (eg, community, hospital) and locations (eg, countries, sites) where the trial was conducted | 6-7 |
| Eligibility criteria | 12a | Eligibility criteria for participants | 6 |
|  | 12b | If applicable, eligibility criteria for sites and for individuals delivering the interventions (eg, surgeons, physiotherapists) | n.a. |
| Intervention and comparator | 13 | Intervention and comparator with sufficient details to allow replication. If relevant, where additional materials describing the intervention and comparator (eg, intervention manual) can be accessed | n.a. |
| Outcomes | 14 | Prespecified primary and secondary outcomes, including the specific measurement variable (eg, systolic blood pressure), analysis metric (eg, change from baseline, final value, time to event), method of aggregation (eg, median, proportion), and time point for each outcome | 7-9 |
| Harms | 15 | How harms were defined and assessed (eg, systematically, non-systematically) | n.a. |
| Sample size | 16a | How sample size was determined, including all assumptions supporting the sample size calculation | 6 |
|  | 16b | Explanation of any interim analyses and stopping guidelines | n.a. |
| **Randomisation:** |  |  |  |
| Sequence generation | 17a | Who generated the random allocation sequence and the method used | n.a. |
|  | 17b | Type of randomisation and details of any restriction (eg, stratification, blocking and block size) | 7 |

|  |  |  | Reported on<br>page no. |
| --- | --- | --- | --- |
| Allocation concealment mechanism | 18 | Mechanism used to implement the random allocation sequence (eg, central computer/telephone; sequentially numbered, opaque, sealed containers), describing any steps to conceal the sequence until interventions were assigned | n.a. |
| Implementation | 19 | Whether the personnel who enrolled and those who assigned participants to the interventions had access to the random allocation sequence | n.a. |
| Blinding | 20a | Who was blinded after assignment to interventions (eg, participants, care providers, outcome assessors, data analysts) | n.a. |
|  | 20b | If blinded, how blinding was achieved and description of the similarity of interventions | n.a. |
| Statistical methods | 21a | Statistical methods used to compare groups for primary and secondary outcomes, including harms | 9-10 |
|  | 21b | Definition of who is included in each analysis (eg, all randomised participants), and in which group | 9-10 |
|  | 21c | How missing data were handled in the analysis | 9 |
|  | 21d | Methods for any additional analyses (eg, subgroup and sensitivity analyses), distinguishing prespecified from post hoc | 10 |
| **Results** |  |  |  |
| Participant flow, including flow diagram | 22a | For each group, the numbers of participants who were randomly assigned, received intended intervention, and were analysed for the primary outcome | 10 |
|  | 22b | For each group, losses and exclusions after randomisation, together with reasons | n.a. |
| Recruitment | 23a | Dates defining the periods of recruitment and follow-up for outcomes of benefits and harms | n.a. |
|  | 23b | If relevant, why the trial ended or was stopped | n.a. |
| Intervention and comparator delivery | 24a | Intervention and comparator as they were actually administered (eg, where appropriate, who delivered the intervention/comparator, how participants adhered, whether they were delivered as intended (fidelity)) | n.a. |
|  | 24b | Concomitant care received during the trial for each group | n.a. |
| Baseline data | 25 | A table showing baseline demographic and clinical characteristics for each group | 10 |
| Numbers analysed, outcomes and estimation | 26 | For each primary and secondary outcome, by group:<br>the number of participants included in the analysis<br>the number of participants with available data at the outcome time point<br>result for each group, and the estimated effect size and its precision (such as 95% confidence interval)<br>for binary outcomes, presentation of both absolute and relative effect size | 10 |
| Harms | 27 | All harms or unintended events in each group | n.a. |
| Ancillary analyses | 28 | Any other analyses performed, including subgroup and sensitivity analyses, distinguishing pre-specified from post hoc | 16 |
| **Discussion** |  |  | 17 |
| Interpretation | 29 | Interpretation consistent with results, balancing benefits and harms, and considering other relevant evidence | 17-20 |
| Limitations | 30 | Trial limitations, addressing sources of potential bias, imprecision, generalisability, and, if relevant, multiplicity of analyses | 20 |

Citation: Hopewell S, Chan AW, Collins GS, Hróbjartsson A, Moher D, Schulz KF, et al. CONSORT 2025 Statement: updated guideline for reporting randomised trials. BMJ. 2025; 388:e081123. https://dx.doi.org/10.1136/bmj-2024-081123
© 2025 Hopewell et al. This is an Open Access article distributed under the terms of the Creative Commons Attribution License (https://creativecommons.org/licenses/by/4.0/), which permits unrestricted use, distribution, and reproduction in any medium, provided the original work is properly cited.
\*We strongly recommend reading this statement in conjunction with the CONSORT 2025 Explanation and Elaboration and/or the CONSORT 2025 Expanded Checklist for important clarifications on all the items. We also recommend reading relevant CONSORT extensions. See www.consort-spirit.org.

## Declaration of interests

☐ The authors declare that they have no known competing financial interests or personal relationships that could have appeared to influence the work reported in this paper.
☒ The authors declare the following financial interests/personal relationships which may be considered as potential competing interests:

Birte Jessen reports financial support was provided by MSH Medical School Hamburg. If there are other authors, they declare that they have no known competing financial interests or personal relationships that could have appeared to influence the work reported in this paper.

## Declaration of interests

Christian Schmidt-Lauber reports financial support was provided by Federal Ministry of Education and Research Berlin Office. If there are other authors, they declare that they have no known competing financial interests or personal relationships that could have appeared to influence the work reported in this paper.

## Declaration of interests

Not correlated to this work: TBH reports consultancy agreements with Alexion, AstraZeneca, Bayer, Beren Therapeutics, Boehringer-Ingelheim, DaVita, Euroimmun, Fresenius Medical Care, Nipoka, Novartis, Otsuka, Pfizer, ProKidney, Protalix, Renovate, Sanofi, Travere, Vera Therapeutics, Vifor, and Vivoryon therapeutics; and research funding from Amicus Therapeutics, Fresenius Medical Care, and Euroimmun. TBH is Co-founder of Global Immune GmbH and holds the patents EP23154267.1, EP23154266.3, EP2015/066881 and 3/2024 PAT 1878 LU. If there are other authors, they declare that they have no known competing financial interests or personal relationships that could have appeared to influence the work reported in this paper.

## Declaration of interests

Bernd Lowe, M.D. reports financial support was provided by University Medical Center Hamburg-Eppendorf, Hamburg, Germany. If there are other authors, they declare that they have no known competing financial interests or personal relationships that could have appeared to influence the work reported in this paper.

## Declaration of interests

☒ The authors declare that they have no known competing financial interests or personal relationships that could have appeared to influence the work reported in this paper.
☐ The authors declare the following financial interests/personal relationships which may be considered as potential competing interests:

## Declaration of interests

Meike Shedden-Mora, Tobias B. Huber reports financial support was provided by German Research Foundation. Meike Shedden-Mora reports a relationship with German Research Foundation that includes: funding grants. Meike Shedden-Mora reports a relationship with German Academic Exchange Service that includes: funding grants and travel reimbursement. Meike Shedden-Mora reports a relationship with Norddeutscher Rundfunk that includes: speaking and lecture fees. Meike Shedden-Mora reports a relationship with European Association of Psychosomatic Medicine (EAPM) that includes: board membership. Meike Shedden-Mora reports a relationship with PKD Cure e.V. that includes: board membership. If there are other authors, they declare that they have no known competing financial interests or personal relationships that could have appeared to influence the work reported in this paper.

## Notes

### Competing Interest Statement

The authors have declared no competing interest.

### Author Declarations

The study protocol was approved by the ethics committee of the State of Hamburg Chamber of Medical Practitioners (reference number 2020-10195-BO-ff) and was preregistered on ISRCTN (ISRTCN16137374).

